# Predictive modeling of morbidity and mortality in COVID-19 hospitalized patients and its clinical implications

**DOI:** 10.1101/2020.12.02.20235879

**Authors:** Joshua M. Wang, Wenke Liu, Xiaoshan Chen, Michael P. McRae, John T. McDevitt, David Fenyö

## Abstract

Clinical activity of 3740 de-identified COVID-19 positive patients treated at NYU Langone Health (NYULH) were collected between January and August 2020. XGBoost model trained on clinical data from the final 24 hours excelled at predicting mortality (AUC=0.92, specificity=86% and sensitivity=85%). Respiration rate was the most important feature, followed by SpO2 and age 75+. Performance of this model to predict the deceased outcome extended 5 days prior with AUC=0.81, specificity=70%, sensitivity=75%. When only using clinical data from the first 24 hours, AUCs of 0.79, 0.80, and 0.77 were obtained for deceased, ventilated, or ICU admitted, respectively. Although respiration rate and SpO2 levels offered the highest feature importance, other canonical markers including diabetic history, age and temperature offered minimal gain. When lab values were incorporated, prediction of mortality benefited the most from blood urea nitrogen (BUN) and lactate dehydrogenase (LDH). Features predictive of morbidity included LDH, calcium, glucose, and C-reactive protein (CRP). Together this work summarizes efforts to systematically examine the importance of a wide range of features across different endpoint outcomes and at different hospitalization time points.

## INTRODUCTION

The first cluster of SARS-CoV-2 was reported in Wuhan, Hubei Province on December 31, 2019. Inciting symptoms remarkably similar to pneumonia, the disease quickly traveled around the world, earning its pandemic status by the World Health Organization on March 11, 2020. Although the first wave has since passed for hardest-hit regions such as New York City (NYC) and most of Asia, a resurgence of cases has already been reported in Europe and record new cases tallied in the Midwest and rural United States (US). As of November 12^th^, the US alone logged its highest tally to date with a 317% growth over the preceding 30 days^1^. The coronavirus disease (COVID-19) is far from seeing the end of its days and there remains a compelling need to prioritize care and resources for patients at elevated risk of morbidity and mortality.

Previous work building machine learning models used patient data from Tongji Hospital^2^,3 (Wuhan, China), Zhongnan Hospital^4^ (Wuhan China), Mount Sinai Hospital^5^ (NYC, US), and NYU Family Health Center^6^ (NYC, US). Surprisingly, clinical features selected varied widely across studies. For example, while McRae et al.’s 2-tiered model^6^ trained on 701 NYC patients to predict mortality was based on actual age, C-reactive protein (CRP), procalcitonin, and D-dimer, Yan et al.’s model^2^ trained on 485 patients from Wuhan selected lactate dehydrogenase (LDH), lymphocyte count, and CRP as the most predictive for mortality. Variations in selected features differed greatly even when trained to predict similar outcomes on data from patients of the same city. Yao et al.’s model^3^ was trained on 137 patients from Wuhan and relied on 28 biomarkers in their final model to predict morbidity. Given the differences among prior models, some of which were driven by domain-specific knowledge, we decided to systematically examine the importance of a wide range of features across different endpoint outcomes and at different hospitalization time points.

This study analyzes retrospective PCR-confirmed COVID-19 inpatient data collected at NYU Langone Hospital spanning 1/1/2020 to 8/7/2020 to predict three sets of clinical outcomes: alive vs deceased, ventilated vs not ventilated, or ICU admitted vs not ICU admitted. The clinical information of 3,740 patient encounters included demographic data (age, sex, insurance, past diagnosis of diabetes, presence of cardiovascular comorbidities), vital signs (SpO2, pulse, respiration rate, temperature, systolic blood pressure, diastolic blood pressure), and the 50 most frequently ordered lab tests in our dataset. Models were developed using two methods: logistic regression with feature selection using Least Absolute Shrinkage and Selection Operator^7^ (LASSO) and gradient tree boosting with XGBoost^8^. An explainable algorithm, such as logistic regression, provides easy to interpret insights into the features of importance. Conversely, the larger model capacity of XGBoost better handles data complexities to explore the extent that predictive performance can be optimized. Together, these methods ensure a holistic survey that explores the clinical underpinnings of disease etiology and the prospects of building models that are sufficiently competent to be effective decision support tools.

## METHODS

### Ethics Statement

Ethics exemption/waiver was confirmed through the Institutional Review Board at NYU Grossman School of Medicine. An IRB self-certification form was completed to ensure that the subsequent research did not fall under human subject research, and so no IRB approval is required. The COVID-19 De-identified Clinical Database was stripped of all unique identifiers prior to receiving data. In addition, all dates were shifted by an arbitrary number of days for each patient. These safeguards ensure that patient data cannot be re-identified, and thus are not subject to HIPAA restrictions on research use, and do not require IRB approval.

### Data Collection

Clinical activity of patients at NYULH was obtained from Epic between 1/1/2020 and 8/7/2020. The data was stripped of all unique identifiers (MRN, names, etc.) and actual dates were shifted by an arbitrary number of days for each patient, which ensures that no data is subject to HIPAA restrictions and thus does not require IRB approval.

### Clinical Data Pre-processing and Cleaning

Our dataset contained 206,677 patients who were tested for COVID-19, of which 12,473 tested positive. Not all patients who tested positive sought hospital care, and without vital signs or lab values, these patients were excluded from analysis. In addition, a majority of the 175,507 patients diagnosed with COVID-19 did not receive in-house PCR tests, which makes it difficult to distinguish which hospital encounters were related to seeking COVID-19 treatment. Thus, only patients for which we could confirm a positive PCR test as reported by NYULH were included. The timestamp of the first encounter in which a PCR test returned positive was used as the starting date for each patient, and the ending date as either the time of discharge for that encounter or time of death. The clinical features that were collected for each patient along with their definitions and additional processing steps are described as follows:

#### Categorical features

- Binned ages: To comply with HIPAA restrictions on research use, exact patient ages were removed and binned into predefined ranges, as determined by the Data Handling Committee.
  - 0-17
  - 18-44
  - 45-64
  - 65-74
  - 75+
- Gender:
  - 0 for Female
  - 1 for Male
- Insurance type:
  - 0 for PPO
  - 1 for EPO, HMO, POS, Indemnity, Medicare, Medicare Managed Care, No Fault, Workers’ Compensation
  - 2 for Medicaid, Medicaid Managed Care
- Diabetes:
  - 1 for any past diagnosis mentioning diabetes
  - 0 otherwise
- Cardiovascular Comorbidities:
  - 1 for any of the following ICD-10 diagnosis codes: I10-I16 (hypertensive diseases), I20-I25 (ischemic heart diseases), I50 (heart failure), I60-I69 (cerebrovascular diseases), and I72 (other aneurysms)
  - 0 otherwise

#### Continuous Features

For each of the following continuous features, multiple periodic measurements were recorded for each patient by vital signs monitors. Measurements were binned into 24 hour windows that began from time of hospitalization. Within each window, values were averaged. Values were then standardized to a mean of zero and variance of one. For each day, encounters without all features listed below were removed and not imputed.

- SpO2 (%)
- Pulse (bpm)
- Respiration Rate (bpm)
- Temperature (°F)
- Systolic Blood Pressure (mmHg)
- Diastolic Blood Pressure (mmHg)

#### Outcomes

- Living status:
  - 0 for Alive
  - 1 for Dead
- Ventilation at any point during hospitalization:
  - 0 for No (did not receive any form of ventilation or only received non-invasive treatments (ex. nasal cannula, non-rebreather mask etc.)
  - 1 for Yes (received mechanical ventilation treatment)
- ICU admission for any duration during hospitalization: Criteria determined by medical triage team, balanced between disease severity and hospital resource availability.
  - 0 for No
  - 1 for Yes

### Lab Data Selection and Cleaning

Lab tests with at least 50% completeness during the first 24 hours for all encounters were considered. Of the 54 lab tests meeting these requirements, EGFR (non-African and African American) was removed due to the formula’s dependency on lab features already selected (creatinine). In addition, the placeholder for ordering a CBC with Differential test and COVID PCR tests were also removed. Missing lab values were imputed using the multivariate imputation by chained equations (MICE) algorithm. Five imputations were generated using predictive mean matching. After imputation, lab values were shifted up by one and log transformed. Model-building approaches that incorporated lab features had individual models built for each imputation.

### Feature Selection and Model Building

All models were trained with a train:validation split of 90:10. Features for logistic regression were selected using Least Absolute Shrinkage and Selection Operator (LASSO) and optimized for a penalty parameter that was one standard error above the minimum deviance for additional shrinkage. The XGBoost parameters were identified using a hyper-parameter search within the following constraints: nrounds: 1000, eta: 0.3, 0.1, 0.01, max_depth = 2, 3, 4, 5, 6, 7, 8, min_child_weight = 0 to 1 by 0.1 increments and gamma = 0 to 1 by 0.1 increments.

For models that were trained on the final day of discharge/death, the performance on predicting outcomes in all preceding days was evaluated on the entire dataset rather than just a 10% subset. Data from previous days was not used in the training of these endpoint models, and thus can all serve as validation.

### Time Series Modeling

In each feature setting, all variables were combined and missing values at each time point were imputed with the immediate previous value (forward filling). After imputation, time points with incomplete feature measurements were discarded, and each patient record was segmented into non-overlapping sequences of length 8. Patients were randomly assigned to training, validation and testing groups in an 8:1:1 ratio for three independent splits. All models were implemented in Python with built-in units in TensorFlow 2 and Keras^9^. Logistic regression was fit as a neural network with the sigmoid output node immediately after the input layer. For MLP, RNN, GRU and LSTM models, a hidden layer of size 8 was added, and the time series models (RNN, GRU and LSTM) were unrolled over 8 time points and trained with true labels provided at each step. Five randomly initialized models were trained for all architectures on each training/validation/testing split. Model performance was evaluated based on all single time point predictions and reported as mean value across all splits.

## RESULTS

More than half of all patients in our dataset were over the age of 65, with pediatric patients (0-17) having the lowest representation (Fig. 1A, Supplemental Table S1). Generally, the proportion of deceased patients increased with age, peaking at 38% for 75+, 16% for 45-64, and 0% for pediatric patients. Most patients who were either ventilated or admitted to the ICU belonged to the 65-74 age group followed by 45-64 and 75+.

**Figure 1.**
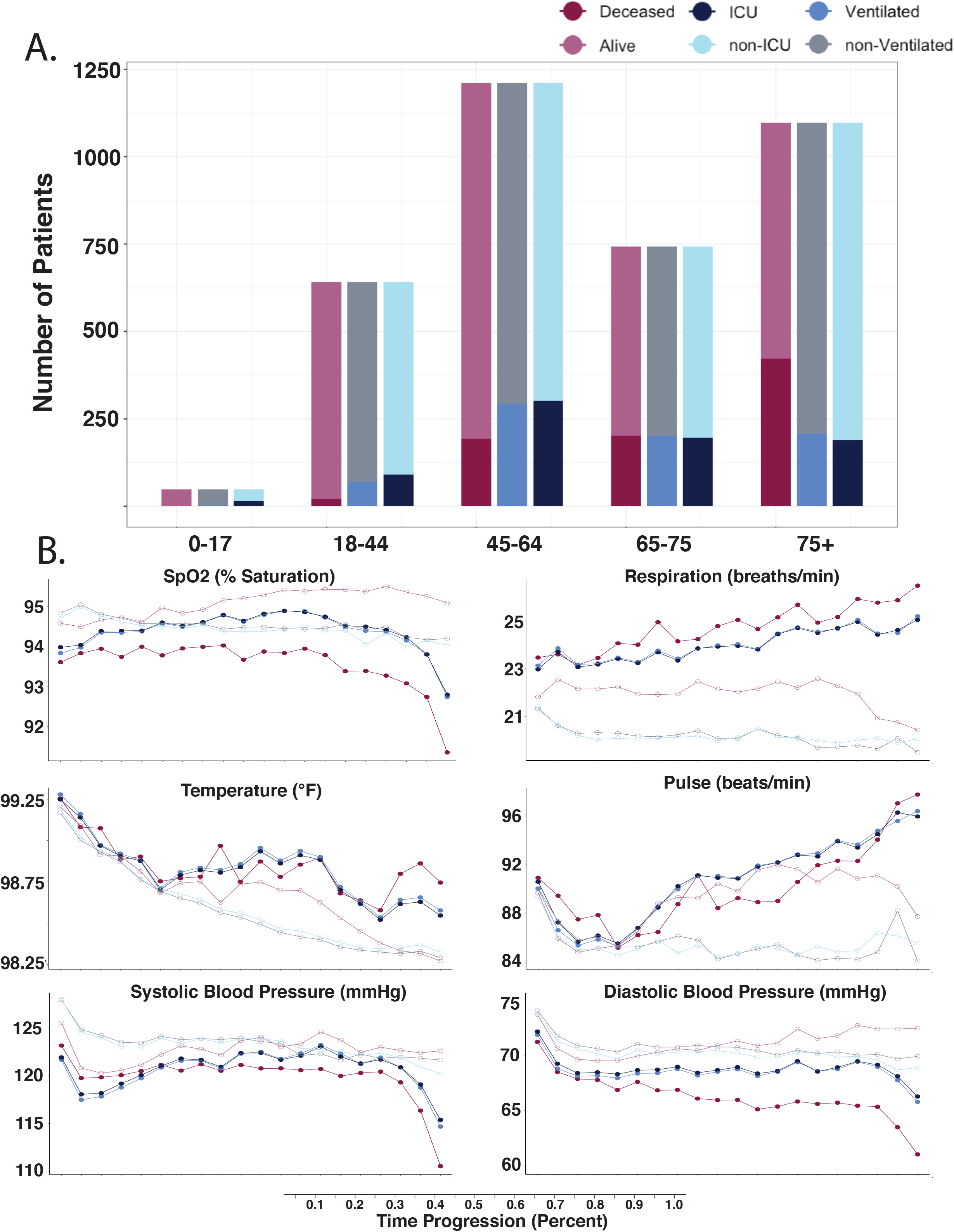
Overview of clinical dataset. **A**. Patient ages were binned by predefined ranges and the ratio of outcomes compared across age groups. **B**. For each patient, hospitalization stay was normalized by length of stay and segmented into 5% windows. Within each window, all values for the measured clinical variable were averaged. Each line is colored by the 6 possible outcomes.

Aggregation of values for commonly acquired clinical metrics over normalized time courses offered meaningful insights into disease progression. Each patient’s hospitalization stay was segmented into 5% windows and clinical metrics were averaged within each bin (Fig. 1B). We first examined the difference of average vital sign measurements between cohorts with different outcomes. The value of SpO2 was statistically different for all three outcome comparisons in the first 5% of hospitalization time (Wilcoxon test; p<2.2e-16, p<2.2e-16, p<2.2e-16). Over the clinical time course, the difference in SpO2 averages increased the most for those that deceased, followed by ICU admitted and ventilated. Differences in respiration rate followed a similar adverse trend with breaths/min increasing the most for those that deceased, followed by ventilated and ICU admitted. The divergence was present even after accounting for overlapping deceased patients. When subsetted for only those that survived, ventilated patients had 2.91 more breaths/min (Wilcoxon test; p<2.2e-16), and ICU admitted patients had 2.90 more breaths/min (Wilcoxon test; p<2.2e-16). At beginning of time course, differences in temperature were small (0.05°F, 0.11°F, and 0.06°F respectively), and not statistically significant for those deceased (Wilcoxon test; p=0.13), but was for those ventilated (Wilcoxon test; p=3.13e-10) or admitted to ICU (Wilcoxon test; p=5.64e-5). Pulse differences at beginning were not significantly different for those ventilated (Wilcoxon test; p=0.29), but was for those deceased (Wilcoxon test; p=7.61e-5) and ICU admitted (Wilcoxon test; p=1.29e-4). Systolic and diastolic blood pressures were continuously lower for patients with worse outcomes in this dataset.

To assess the effectiveness of these vital signs to triage clinical outcomes, only data collected in the first 24 hours after admission was initially considered. Specifically for the ventilation outcome, respiration rates and SpO2 levels may be influenced by when mechanical ventilation was administered. Of 3740 encounters, 7% were ventilated within the first 24 hours of admission. To assess the bias that early administration of mechanical ventilation during the first 24 hours may have on respiration rate and SpO2 levels, the distribution of values was compared against a filtered subset containing only values recorded prior to start of ventilation. At the per-encounter level, the difference in respiration rates (Wilcoxon test; p=0.26, Supplemental Fig. 1A) and SpO2 levels (Wilcoxon test; p=0.20, Supplemental Fig. 1B) were not significantly different. Given that 1) 93% of encounters were not influenced by early ventilation treatment, 2) the insignificant difference in distributions, and 3) the desire to keep feature selection consistent across models, all values recorded within the first 24 hours were included. For each encounter, continuous features with multiple recordings (SpO2, pulse, respiration rate, temperature, systolic blood pressure, diastolic blood pressure) were averaged, and then standardized to mean of 0 and standard deviation of 1.

For logistic regression, features were selected using LASSO with 10-fold cross validation. Grid search was used to optimize XGBoost parameters (Supplemental Table S2). When trained on data from the first 24 hours, the logistic model had AUC performances of 0.79, 0.80, 0.77, specificities of 59%, 78%, 79%, and sensitivities of 86%, 74%, 68% respectively (Fig. 2A). XGBoost performed similarly with AUC performances of 0.80, 0.80, 0.77, specificities of 59%, 83%, 69%, and sensitivities of 86%, 70%, 77% respectively (Fig. 2B).

**Figure 2.**
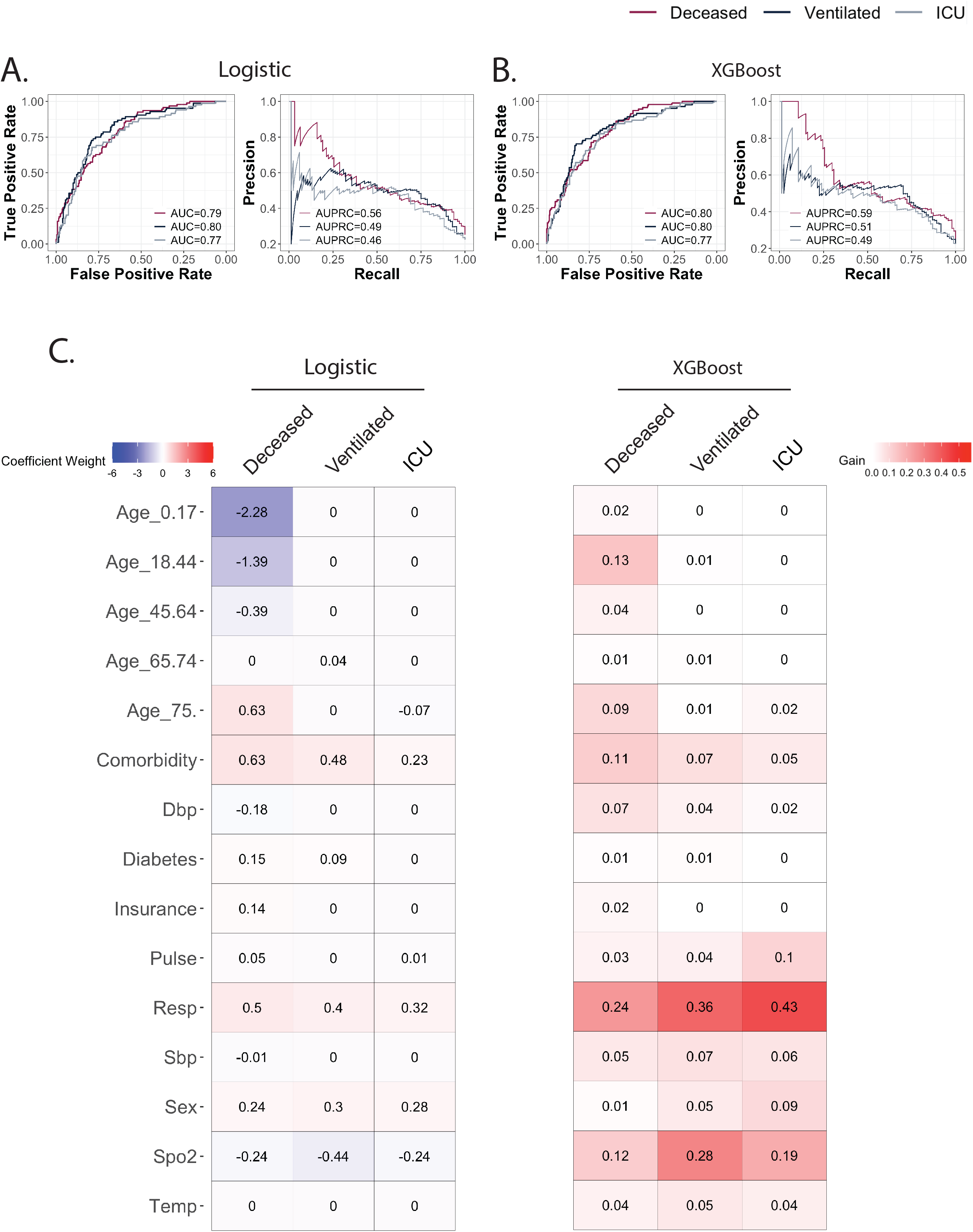
Predictive performance using clinical data from the first 24 hours. **A**. ROC curve and PRC for logistic regression model. **B**. ROC curve and PRC for XGBoost model. **C**. Coefficient weights for the logistic model are recorded on the left. Model performance gains for XGBoost are listed on the right.

In both logistic regression and gradient tree boosting settings, features of importance varied across clinical outcomes (Fig. 2C). For logistic regression models of the three outcomes, respiration rate, SpO2 and cardiovascular comorbidity were among predictive features, but age groups were selected only for predicting mortality. For boosting tree models, feature importance measures showed that respiration rate was consistently the most important feature for all three outcomes, and age_18-44 was the second most important feature only for vital status. Respiration rate and SpO2 were important for predicting all three outcomes. Differences in temperature were not strongly predictive in any cohort in either model, and its insignificant difference in the deceased outcome group together suggests that its role in screening for increased disease severity may not be dependable.

The 50 most frequently collected labs and their relative importance were also studied. A t-SNE plot (Fig. 3A) suggests lack of clustering among lab features, and overall low correlation (Fig. 3B) in pairwise comparisons (|μ| = 0.08, |σ| = 0.10). Local pockets of correlation (|cor|>=0.83) were identified between (hemoglobin, hematocrit, red blood cell count), (absolute neutrophils, white blood cell count), and (bilirubin direct, bilirubin total). Each of these sets measures variables that are clinically interdependent and thus expected.

**Figure 3.**
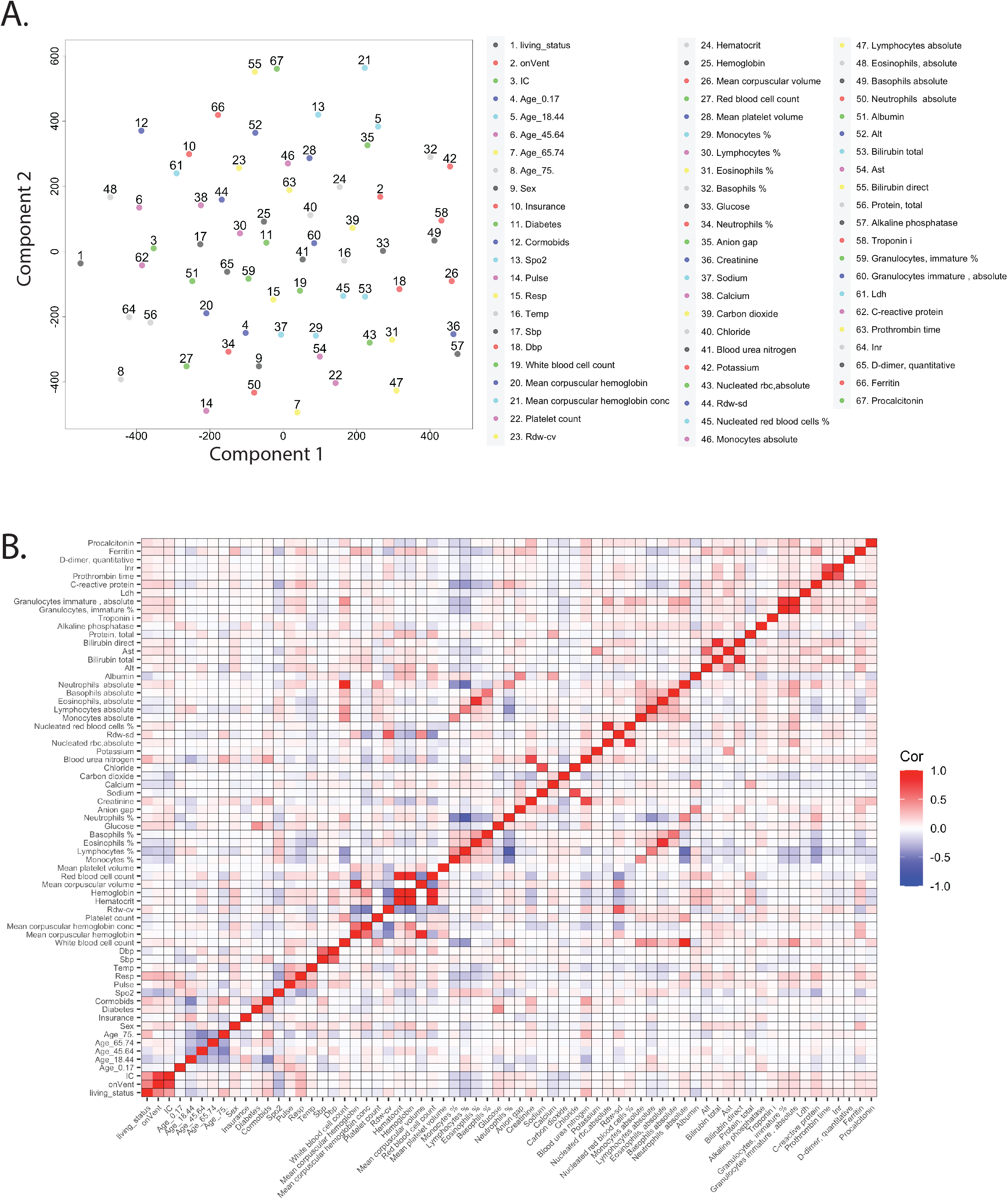
Overview of lab features collected in the first 24 hours. **A**. t-SNE plot based on previously collected clinical features and new lab values. **B**. Pairwise Pearson correlation heatmap.

Incorporating lab features into the predictive models marginally improved performance. Logistic regression had AUC performances of 0.83, 0.81, 0.78, specificities of 68%, 70%, 69%, and sensitivities of 85%, 83%, 74% respectively (Fig. 4A). The XGBoost model performed better with AUC increasing to 0.84, 0.79, 0.78, specificities of 71%, 72%, 65%, and sensitivities of 83%, 73%, 78% respectively (Fig. 4B). For logistic regression, blood urea nitrogen (BUN) and albumin were among the lab features (Fig. 4C) predictive of mortality. The XGBoost model found most performance gain from BUN and LDH. Feature importance for predicting ventilation or ICU admission differed between models. For ventilation, logistic regression selected calcium, glucose, and CRP with large absolute coefficient values, while XGBoost identified calcium, glucose, CRP, and LDH as important features. For those admitted to ICU, XGBoost benefited from the same lab features, while monocyte percentage and carbon dioxide were additionally selected for by logistic regression. Of note, for XGBoost, no lab feature showed higher importance measure than respiration rate and SpO2 did for all three outcomes.

**Figure 4.**
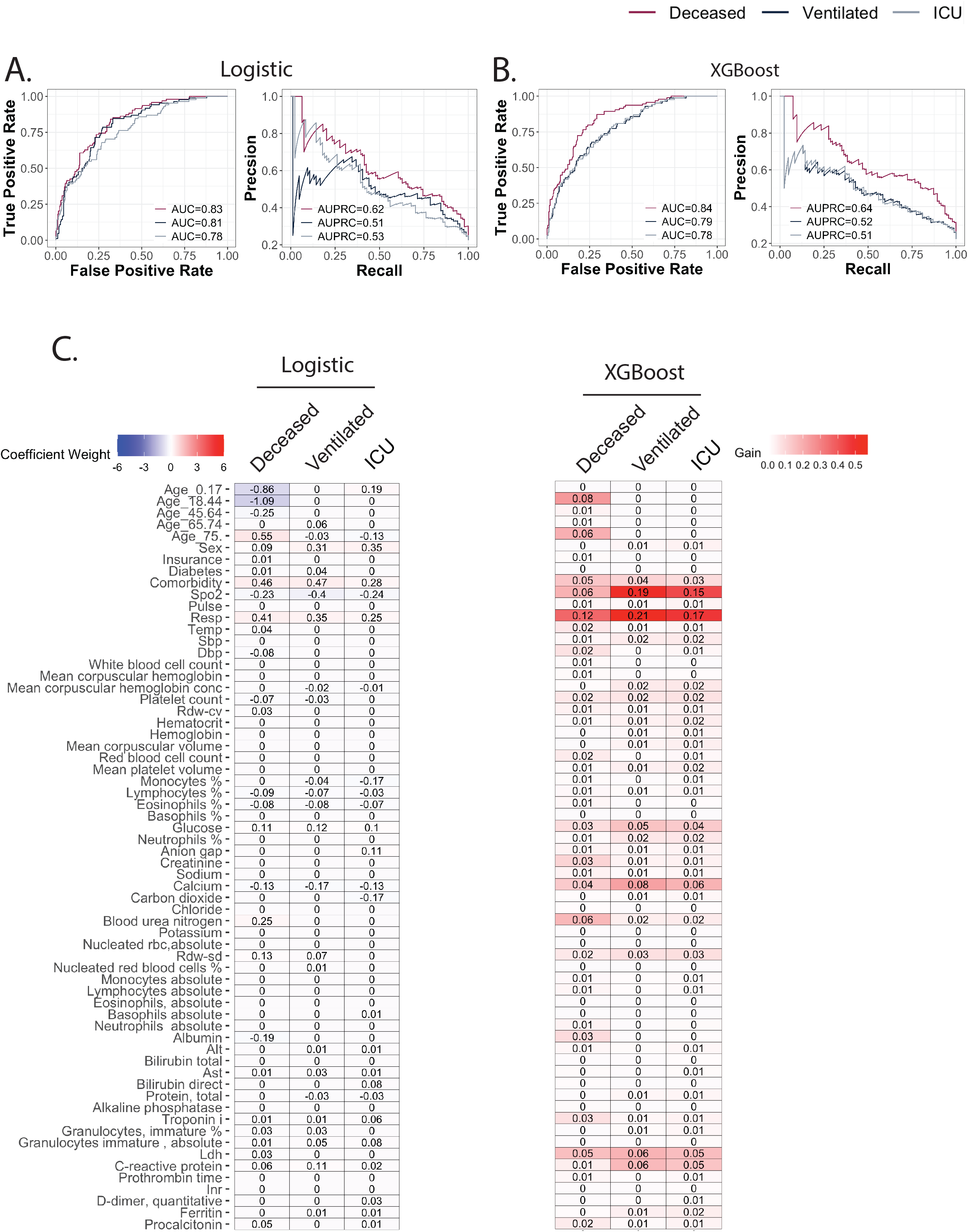
Predictive performance after incorporating lab features. **A**. ROC curve and PRC for logistic regression model. **B**. ROC curve and PRC for XGBoost model. **C**. Coefficient weights for the logistic model are recorded on the left. Model performance gains for XGBoost are listed on the right.

Finally, models trained on data collected in the last 24 hours excelled at predicting deceased. The logistic regression model (Fig. 5A) had AUC performance of 0.91, specificity of 88% and sensitivity of 84%. The XGBoost model (Fig. 5B) had AUC performance of 0.92% specificity of 86% and sensitivity of 85%. The importance of respiration rate increased for XGBoost (Fig. 5C), accounting for more than 50% of the gain. Values of SpO2 and age 75+ were the next most important features.

**Figure 5.**
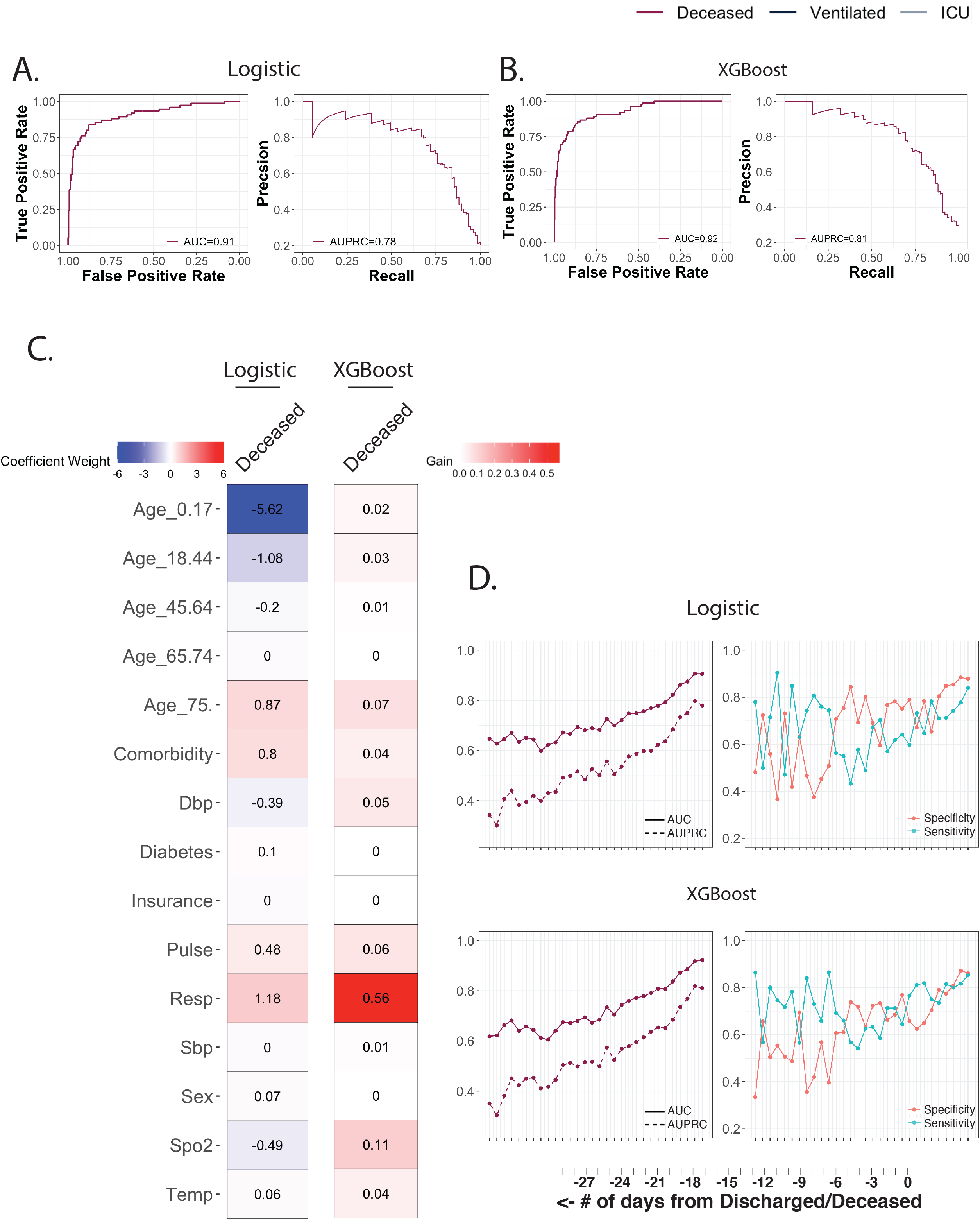
Predictive performance of deceased using clinical data from the final 24 hours. **A**. ROC curve and PRC for logistic regression model. **B**. ROC curve and PRC for XGBoost model. **C**. Coefficient weights for the logistic model are recorded on the left. Model performance gains for XGBoost are listed on the right. **D**. Performance of models to predict deceased outcome was assessed using clinical data from the preceding 30 days. Plots track the AUC, AUPRC, specificity, and sensitivity when using the threshold that maximized the sum of the sensitivity and specificity (Youden’s J statistic).

Using the same coefficients and tree weights/structures, both models were assessed based on clinical data from the preceding 30 days (Fig. 5D). With cutoffs of 0.80 for AUC, and 70% for specificity and sensitivity, logistic regression was able to predict deceased 4 days in advance (AUC = 0.82, specificity = 85%, sensitivity = 71%) and 5 days in advance (AUC = 0.81, specificity = 70%, sensitivity = 75%) for XGBoost. Models were not trained on those ventilated or ICU admitted, as these events are unlikely to occur in the final 24 hours preceding discharge/deceased. Labs were not incorporated because few blood tests were ordered in the final 24 hours.

To explore whether patient status can be dynamically predicted based on history data, we also built time series models using simple recurrent neural network (RNN), gated recurrent unit (GRU) and long short term memory (LSTM) architectures and compared the performance metrics to single time point models of logistic regression (LR) and multilayer perceptron (MLP). The vital status of each patient was converted to a time series that flagged positive for time points within 3-day intervals before the patient deceased. Model comparison was carried out with three different feature sets: vital signs (body temperature, pulse, respiration rate, systolic blood pressure, diastolic blood pressure, SpO2) only, vital signs and 46 lab results with nonzero coefficients in the single time point LASSO regression model, vital signs and lab results plus ‘static’ demographical information of sex, age group, diabetic history and comorbidities (Supplemental Table S3). As the time series data were recorded with uneven and irregular intervals, the progression time (in days) was included in all models as an additional feature. For models only including vital sign features, time series models showed better performance (Fig. 6) compared to single time point models, but performance was comparable among all models when lab results and demographical information was added to the feature set.

**Figure 6.**
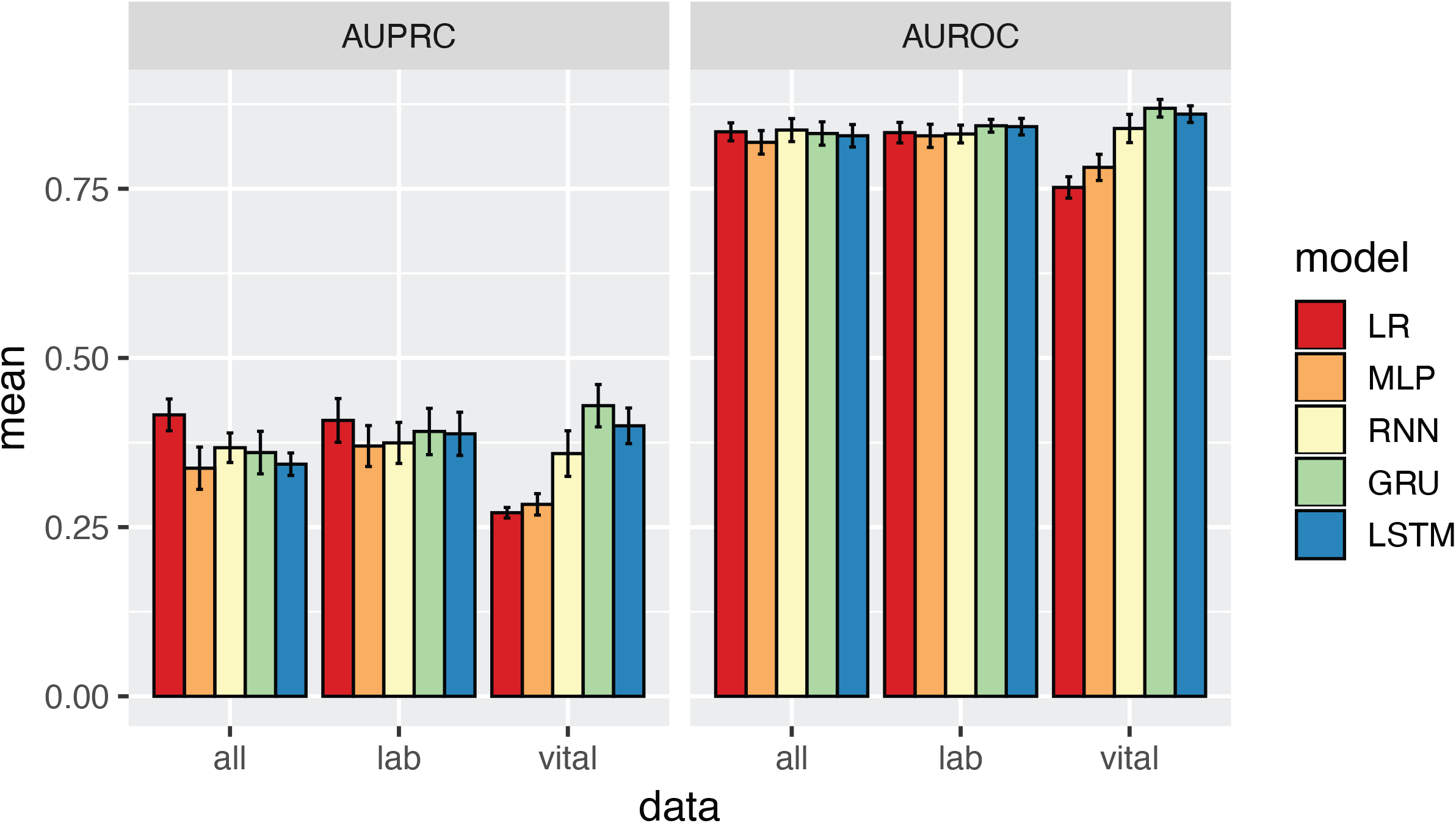
Time series model performance. Mean values of area under the precision-recall curve (AUPRC) and area under the receiver operating characteristics curve (AUROC) for five model architectures across three feature settings. Vital: only include progression time and vital signs. Lab: all vital sign features and 46 laboratory results. All: vital sign and laboratory variables and static demographic features.

## DISCUSSION

Retrospective analysis of COVID-19 positive patients identified recognizable clinical markers such as respiration rate and SpO2, but also provided insights distinguishing morbidity (ICU admitted or ventilated outcomes) from mortality (deceased outcome). Our results aligned with previous work^10^ analyzing patient data from NYU Langone to predict absence of adverse events within a 96 hour window as opposed to negative outcomes. Several features of importance overlapped both studies, notably respiration rate, SpO2, LDH, BUN, and CRP. However, other selected features such as temperature, platelet count, pulse, and eosinophil percentage were found to not be important in our model.

Although the goal of stratifying patients by disease severity aligned, the different approaches likely explain the differences in variable explanation. Our study differs in that our models are trained only on clinical data from the first 24 hours after admission, as compared to continuously updating predictions when new labs are reported. Thus, features that are important for outcome prediction at time of admission will differ from those that better model variations in disease severity over time. In addition, we stratify our negative outcomes into mortality and morbidity, and separate morbidity further to compare those requiring ICU admission versus ventilation. Eosinophil percentage was statistically different between all 3 clinical outcomes, while temperature and pulse were only different for morbidity and platelet counts only for mortality (Supplemental Table S1). It is hypothesized that patients exhibiting symptoms of fever and increased pulse rate, likely a consequence of decreased SpO2 (cor = −0.21, −0.12 respectively) will likely be prioritized for ICU care and/or ventilation. Although SpO2 and respiration rate were consistently selected as predictive features across outcomes and modeling methods, age groups were informative predictors of mortality risk only. As expected, the mortality model performed better than morbidity models. These results suggest that disease severity and mortality risks may require unique modeling with different predictor subsets and weighting factors. It is also consistent with the observation that senior patients were the most vulnerable population, while mortality rate among the youth was relatively low^11^.

In addition, although current evidence suggests adults with type 2 diabetes mellitus are at increased risk for COVID-19 complications, our XGBoost model did not find a past diagnosis important for predicting morbidity or mortality. Only after incorporating lab features did we identify a positive correlation between exact glucose values and poorer outcomes. Together this observation suggests that the elevated blood sugar levels observed may be the result of physiological stress triggered by the disease. Indeed, prior work has shown that even when controlled for pre-existing diabetes, hyperglycemia was commonly observed in acutely ill hospitalized patients and linked to poorer outcomes^12,13^.

Other lab features also identified routine chemistry data points that shed light on disease pathology. Values of LDH were elevated for all three clinical outcomes, a finding consistent with widespread tissue damage that has been shown in numerous studies to be a predictor of morbidity and mortality in a wide variety of diseases beyond COVID-19^14–18^. Mortality was also predicted for by BUN. To investigate further the possibility of any relations to acute kidney injury, we re-trained our models with BUN/creatinine ratio as an additional feature. While correlated with mortality (cor=0.17), the feature was not selected for by LASSO, and was only of importance when BUN was removed from training. Indeed, recent literature has revealed that BUN is emerging as an independent predictor of mortality in a variety of diseases, including heart failure^19^, aortic dissection^20^, and acute pancreatitis^21^. It has also been proposed that BUN is an important indicator for metabolic diseases and general nutritional status of patients, explaining its relative importance in the prediction for mortality. The relationship here is unclear and warrants further investigation.

Interestingly, admission calcium level was a more important predictor of morbidity in our models than procalcitonin was. As a peptide precursor of calcitonin, a hormone involved in calcium homeostasis, procalcitonin is also an acute phase reactant that has been used historically (albeit controversially) to help diagnose bacterial pneumonia^22–24^. Although many studies^25–27^ have described a positive relationship between procalcitonin levels and mortality and morbidity in COVID-19 patients, few have commented on the importance of calcium as a prognostic value, as we have found in our study. Calcium was negatively correlated with all 3 measured clinical outcomes, which is consistent with other studies linking hypocalcemia with increased morbidity and mortality in COVID-19 patients^28–30^. Theoretically, hypocalcemia could be a result of increased procalcitonin, since procalcitonin is the precursor of calcitonin whose function is to reduce serum calcium. Interestingly, it has been reported that in a systemic inflammatory response, serum calcitonin does not increase concordantly in response to increased procalcitonin. This situation could indicate that calcium is a predictive factor through an entirely different mechanism than the more well-established procalcitonin. One theory is that alteration of calcium homeostasis is perhaps used as a strategy by the SARS-CoV-2 virus for survival and replication since calcium is essential for virus structure formation, entry, gene expression, virion maturation and release. Another possibility is that patients who present with hypocalcemia have preexisting parathyroid hormone (PTH) and vitamin D imbalances that are exacerbated by SARS-CoV-2 infection. Our study could not evaluate the importance of PTH or vitamin D due to infrequent lab orders (0.21% and 0.08% completeness respectively).

While the inclusion of lab features resulted in only modest improvement for ventilation and ICU admission prediction, lab values did result in larger increases in performance metrics for mortality prediction. However, time series modeling failed to improve prediction performance with more clinical features. This observation is likely due to the fact that laboratory results were sampled much less frequently than vital sign readings. As data was retrospectively gathered from Epic during the early stages of the pandemic when diagnostic and treatment protocols were still being developed, a concerted effort to gather novel biomarker tests that have later been shown to be linked with disease severity is not expected. Moreover, treating ‘static’ demographical as repeating time series measurements may be suboptimal for recurrent models. As discussed above, laboratory measurements may help modeling mortality risk of patients, and future work will focus on efficiently incorporating these static features for dynamic predictions^31,32^.

## Data Availability

Institutional policies prevent public distribution of patient clinical data.

## DATA AVAILABILITY

The data that support the findings of this study were obtained from the Medical Center Information Technology (MCIT) at NYU Langone Health but restrictions apply to the availability of these data and so are not publicly available due to specific institutional requirements.

## ACKNOWLEDGEMENTS

We wish to thank MCIT and Office of Science & Research at NYU Langone Health for maintaining and de-identifying the clinical database. JMW is supported by the New York University Medical Scientist Training Program (T32GM136573). The content is solely the responsibility of the authors and does not necessarily represent the official views of the National Institutes of Health. A portion of this work was funded by Renaissance Health Service Corporation and Delta Dental of Michigan.

## AUTHOR CONTRIBUTIONS

J.M. Wang, W. Liu, and D. Fenyö contributed to conception and design, data acquisition, analysis, interpretation, drafted and critically revised the manuscript.

X. Chen contributed to conception and design, analysis, interpretation, drafted and critically revised the manuscript.

M.P. McRae, and J.T. McDevitt contributed to conception and design, interpretation, and critically revised the manuscript.

## FIGURE LEGENDS

**Supplemental Figure 1**. Effect of early ventilation treatment on vital signs. A. Distribution of averaged respiration rates for all values preceding 24 hours (top) and for all values that precede initiation of ventilation treatment (bottom). B. Distribution of averaged SpO2 for all values preceding 24 hours (top) and for all values that precede initiation of ventilation treatment (bottom).

