## Supplemental Table S1 for "Predictive modeling of morbidity and mortality in COVID-19 hospitalized patients and its clinical implications"

1. Demographics, vital signs, lab results, and clinical outcomes of retrospective cohort.

|  | **Alive (N2903)** | **Deceased (N837)** | **Total (N3740)** | **p value** |  | **non-Ventilated (N2965)** | **Ventilated (N775)** | **Total (N3740)** | **p value** |  | **ICU (N791)** | **non-ICU (N2949)** | **Total (N3740)** | **p value** |
| --- | --- | --- | --- | --- | --- | --- | --- | --- | --- | --- | --- | --- | --- | --- |
| age_0.17 |  |  |  | 0.01158 |  |  |  |  | 0.818071 |  |  |  |  | 1 |
| 0 | 2855 (98.3%) | 837 (100.0%) | 3692 (98.7%) | |  | 2920 (98.5%) | 772 (99.6%) | 3692 (98.7%) | |  | 776 (98.1%) | 2916 (98.9%) | 3692 (98.7%) | |
| 1 | 48 (1.7%) | 0 (0.0%) | 48 (1.3%) |  |  | 45 (1.5%) | 3 (0.4%) | 48 (1.3%) |  |  | 15 (1.9%) | 33 (1.1%) | 48 (1.3%) |  |
| age_18.44 |  |  |  | < 1e-06 |  |  |  |  | < 1e-06 |  |  |  |  | 0.000082 |
| 0 | 2283 (78.6%) | 816 (97.5%) | 3099 (82.9%) | |  | 2394 (80.7%) | 705 (91.0%) | 3099 (82.9%) | |  | 701 (88.6%) | 2398 (81.3%) | 3099 (82.9%) | |
| 1 | 620 (21.4%) | 21 (2.5%) | 641 (17.1%) |  |  | 571 (19.3%) | 70 (9.0%) | 641 (17.1%) |  |  | 90 (11.4%) | 551 (18.7%) | 641 (17.1%) |  |
| age_45.64 |  |  |  | < 1e-06 |  |  |  |  | 0.018414 |  |  |  |  | 0.007867 |
| 0 | 1885 (64.9%) | 644 (76.9%) | 2529 (67.6%) | |  | 2047 (69.0%) | 482 (62.2%) | 2529 (67.6%) | |  | 490 (61.9%) | 2039 (69.1%) | 2529 (67.6%) | |
| 1 | 1018 (35.1%) | 193 (23.1%) | 1211 (32.4%) | |  | 918 (31.0%) | 293 (37.8%) | 1211 (32.4%) | |  | 301 (38.1%) | 910 (30.9%) | 1211 (32.4%) | |
| age_65.74 |  |  |  | 0.040986 |  |  |  |  | 0.000076 |  |  |  |  | 0.006167 |
| 0 | 2361 (81.3%) | 636 (76.0%) | 2997 (80.1%) | |  | 2424 (81.8%) | 573 (73.9%) | 2997 (80.1%) | |  | 595 (75.2%) | 2402 (81.5%) | 2997 (80.1%) | |
| 1 | 542 (18.7%) | 201 (24.0%) | 743 (19.9%) |  |  | 541 (18.2%) | 202 (26.1%) | 743 (19.9%) |  |  | 196 (24.8%) | 547 (18.5%) | 743 (19.9%) |  |
| age_75. |  |  |  | < 1e-06 |  |  |  |  | 1 |  |  |  |  | 0.009921 |
| 0 | 2228 (76.7%) | 415 (49.6%) | 2643 (70.7%) | |  | 2075 (70.0%) | 568 (73.3%) | 2643 (70.7%) | |  | 602 (76.1%) | 2041 (69.2%) | 2643 (70.7%) | |
| 1 | 675 (23.3%) | 422 (50.4%) | 1097 (29.3%) | |  | 890 (30.0%) | 207 (26.7%) | 1097 (29.3%) | |  | 189 (23.9%) | 908 (30.8%) | 1097 (29.3%) | |
| sex |  |  |  | 0.000491 |  |  |  |  | < 1e-06 |  |  |  |  | < 1e-06 |
| 0 | 1328 (45.7%) | 310 (37.0%) | 1638 (43.8%) | |  | 1396 (47.1%) | 242 (31.2%) | 1638 (43.8%) | |  | 249 (31.5%) | 1389 (47.1%) | 1638 (43.8%) | |
| 1 | 1575 (54.3%) | 527 (63.0%) | 2102 (56.2%) | |  | 1569 (52.9%) | 533 (68.8%) | 2102 (56.2%) | |  | 542 (68.5%) | 1560 (52.9%) | 2102 (56.2%) | |
| insurance |  |  |  | < 1e-06 |  |  |  |  | 1 |  |  |  |  | 1 |
| 0 | 333 (11.5%) | 33 (3.9%) | 366 (9.8%) |  |  | 295 (9.9%) | 71 (9.2%) | 366 (9.8%) |  |  | 78 (9.9%) | 288 (9.8%) | 366 (9.8%) |  |
| 1 | 1762 (60.7%) | 676 (80.8%) | 2438 (65.2%) | |  | 1905 (64.2%) | 533 (68.8%) | 2438 (65.2%) | |  | 521 (65.9%) | 1917 (65.0%) | 2438 (65.2%) | |
| 2 | 808 (27.8%) | 128 (15.3%) | 936 (25.0%) |  |  | 765 (25.8%) | 171 (22.1%) | 936 (25.0%) |  |  | 192 (24.3%) | 744 (25.2%) | 936 (25.0%) |  |
| diabetes |  |  |  | < 1e-06 |  |  |  |  | 0.000003 |  |  |  |  | 0.01034 |
| 0 | 1630 (56.1%) | 370 (44.2%) | 2000 (53.5%) | |  | 1653 (55.8%) | 347 (44.8%) | 2000 (53.5%) | |  | 376 (47.5%) | 1624 (55.1%) | 2000 (53.5%) | |
| 1 | 1273 (43.9%) | 467 (55.8%) | 1740 (46.5%) | |  | 1312 (44.2%) | 428 (55.2%) | 1740 (46.5%) | |  | 415 (52.5%) | 1325 (44.9%) | 1740 (46.5%) | |
| comorbidity |  |  |  | < 1e-06 |  |  |  |  | < 1e-06 |  |  |  |  | < 1e-06 |
| 0 | 1052 (36.2%) | 97 (11.6%) | 1149 (30.7%) | |  | 1010 (34.1%) | 139 (17.9%) | 1149 (30.7%) | |  | 173 (21.9%) | 976 (33.1%) | 1149 (30.7%) | |
| 1 | 1851 (63.8%) | 740 (88.4%) | 2591 (69.3%) | |  | 1955 (65.9%) | 636 (82.1%) | 2591 (69.3%) | |  | 618 (78.1%) | 1973 (66.9%) | 2591 (69.3%) | |
| SpO2 |  |  |  | < 1e-06 |  |  |  |  | < 1e-06 |  |  |  |  | < 1e-06 |
| Mean (SD) | 0.114 (0.934) | -0.395 (1.115) | -0.000 (1.000) | |  | 0.146 (0.916) | -0.560 (1.105) | -0.000 (1.000) | |  | -0.414 (1.114) | 0.111 (0.937) | -0.000 (1.000) | |
| Range | -4.719 - 2.035 | -6.762 - 2.035 | -6.762 - 2.035 | |  | -4.719 - 2.035 | -6.762 - 2.035 | -6.762 - 2.035 | |  | -6.762 - 2.035 | -4.719 - 2.035 | -6.762 - 2.035 | |
| Pulse |  |  |  | 1 |  |  |  |  | 0.000002 |  |  |  |  | < 1e-06 |
| Mean (SD) | -0.018 (0.988) | 0.063 (1.038) | 0.000 (1.000) | |  | -0.047 (0.979) | 0.179 (1.060) | 0.000 (1.000) | |  | 0.207 (1.115) | -0.056 (0.960) | 0.000 (1.000) | |
| Range | -3.411 - 4.946 | -2.365 - 3.565 | -3.411 - 4.946 | |  | -2.99 - 4.946 | -3.411 - 3.908 | -3.411 - 4.946 | |  | -3.411 - 4.218 | -2.99 - 4.946 | -3.411 - 4.946 | |
| Resp |  |  |  | < 1e-06 |  |  |  |  | < 1e-06 |  |  |  |  | < 1e-06 |
| Mean (SD) | -0.140 (0.932) | 0.485 (1.074) | -0.000 (1.000) | |  | -0.143 (0.924) | 0.548 (1.088) | -0.000 (1.000) | |  | 0.450 (1.067) | -0.121 (0.946) | -0.000 (1.000) | |
| Range | -2.163 - 4.646 | -2.363 - 4.396 | -2.363 - 4.646 | |  | -2.363 - 4.646 | -1.683 - 4.396 | -2.363 - 4.646 | |  | -1.987 - 4.396 | -2.363 - 4.646 | -2.363 - 4.646 | |
| Temp |  |  |  | 1 |  |  |  |  | 0.000006 |  |  |  |  | 0.002541 |
| Mean (SD) | -0.014 (0.983) | 0.050 (1.056) | -0.000 (1.000) | |  | -0.041 (0.963) | 0.157 (1.119) | -0.000 (1.000) | |  | 0.109 (1.155) | -0.029 (0.952) | -0.000 (1.000) | |
| Range | -7.636 - 3.978 | -5.064 - 5.583 | -7.636 - 5.583 | |  | -7.636 - 3.598 | -5.064 - 5.583 | -7.636 - 5.583 | |  | -7.636 - 5.583 | -3.539 - 3.598 | -7.636 - 5.583 | |
| SBP |  |  |  | 1 |  |  |  |  | 1 |  |  |  |  | 1 |
| Mean (SD) | 0.012 (0.977) | -0.040 (1.075) | 0.000 (1.000) | |  | 0.009 (0.994) | -0.035 (1.023) | 0.000 (1.000) | |  | -0.032 (1.095) | 0.009 (0.973) | 0.000 (1.000) | |
| Range | -2.637 - 4.857 | -2.941 - 4.802 | -2.941 - 4.857 | |  | -2.941 - 4.857 | -2.62 - 4.802 | -2.941 - 4.857 | |  | -2.941 - 4.857 | -2.62 - 4.793 | -2.941 - 4.857 | |
| DBP |  |  |  | < 1e-06 |  |  |  |  | 1 |  |  |  |  | 1 |
| Mean (SD) | 0.052 (0.981) | -0.182 (1.042) | -0.000 (1.000) | |  | 0.002 (1.001) | -0.007 (0.996) | -0.000 (1.000) | |  | 0.017 (1.104) | -0.005 (0.970) | -0.000 (1.000) | |
| Range | -3.673 - 7.072 | -2.867 - 4.815 | -3.673 - 7.072 | |  | -3.673 - 7.072 | -3.165 - 4.815 | -3.673 - 7.072 | |  | -3.165 - 7.072 | -3.673 - 5.579 | -3.673 - 7.072 | |
| White blood cell count | |  |  | 0.00379 |  |  |  |  | 0.000066 |  |  |  |  | < 1e-06 |
| Mean (SD) | -0.037 (0.876) | 0.129 (1.161) | -0.000 (0.950) | |  | -0.043 (0.912) | 0.164 (1.068) | -0.000 (0.950) | |  | 0.189 (1.058) | -0.051 (0.912) | -0.000 (0.950) | |
| Range | -3.704 - 3.542 | -4.498 - 9.277 | -4.498 - 9.277 | |  | -4.498 - 3.602 | -3.842 - 9.277 | -4.498 - 9.277 | |  | -4.498 - 6.137 | -4.316 - 9.277 | -4.498 - 9.277 | |
| Mean corpuscular hemoglobin | | |  | 1 |  |  |  |  | 1 |  |  |  |  | 1 |
| Mean (SD) | -0.012 (0.976) | 0.043 (0.985) | -0.000 (0.978) | |  | -0.002 (0.987) | 0.007 (0.946) | -0.000 (0.978) | |  | -0.003 (0.985) | 0.001 (0.977) | -0.000 (0.978) | |
| Range | -7.99 - 5.245 | -4.457 - 2.831 | -7.99 - 5.245 | |  | -7.99 - 5.245 | -4.457 - 2.663 | -7.99 - 5.245 | |  | -4.79 - 3.282 | -7.99 - 5.245 | -7.99 - 5.245 | |
| Mean corpuscular hemoglobin conc | | |  | 0.003443 |  |  |  |  | 1 |  |  |  |  | 1 |
| Mean (SD) | 0.037 (0.941) | -0.127 (0.993) | -0.000 (0.956) | |  | 0.005 (0.922) | -0.019 (1.073) | -0.000 (0.956) | |  | -0.008 (1.070) | 0.002 (0.923) | -0.000 (0.956) | |
| Range | -4.698 - 3.007 | -4.499 - 2.61 | -4.698 - 3.007 | |  | -4.698 - 2.951 | -4.499 - 3.007 | -4.698 - 3.007 | |  | -4.499 - 3.007 | -4.698 - 2.951 | -4.698 - 3.007 | |
| Platelet count | |  |  | 0.000004 |  |  |  |  | 0.31893 |  |  |  |  | 1 |
| Mean (SD) | 0.045 (0.948) | -0.157 (1.019) | -0.000 (0.968) | |  | 0.020 (0.980) | -0.078 (0.916) | -0.000 (0.968) | |  | -0.035 (0.930) | 0.009 (0.978) | -0.000 (0.968) | |
| Range | -11.22 - 2.933 | -5.661 - 2.892 | -11.22 - 2.933 | |  | -11.22 - 2.933 | -5.131 - 2.892 | -11.22 - 2.933 | |  | -5.131 - 2.892 | -11.22 - 2.933 | -11.22 - 2.933 | |
| Rdw-cv |  |  |  | < 1e-06 |  |  |  |  | 1 |  |  |  |  | 1 |
| Mean (SD) | -0.063 (0.969) | 0.218 (0.984) | 0.000 (0.979) | |  | -0.010 (0.982) | 0.037 (0.967) | 0.000 (0.979) | |  | 0.025 (0.977) | -0.007 (0.980) | 0.000 (0.979) | |
| Range | -20.797 - 5.624 | -1.506 - 5.139 | -20.797 - 5.624 | |  | -20.797 - 5.624 | -1.696 - 5.139 | -20.797 - 5.624 | |  | -1.696 - 5.139 | -20.797 - 5.624 | -20.797 - 5.624 | |
| Hematocrit |  |  |  | 1 |  |  |  |  | 0.000811 |  |  |  |  | 0.001182 |
| Mean (SD) | 0.022 (0.943) | -0.075 (1.115) | -0.000 (0.985) | |  | -0.029 (0.978) | 0.111 (1.005) | -0.000 (0.985) | |  | 0.120 (0.990) | -0.032 (0.981) | -0.000 (0.985) | |
| Range | -6.721 - 2.552 | -8.51 - 2.754 | -8.51 - 2.754 | |  | -8.51 - 2.754 | -5.046 - 2.583 | -8.51 - 2.754 | |  | -5.491 - 2.583 | -8.51 - 2.754 | -8.51 - 2.754 | |
| Hemoglobin |  |  |  | 0.433063 |  |  |  |  | 0.002109 |  |  |  |  | 0.002654 |
| Mean (SD) | 0.029 (0.943) | -0.101 (1.095) | 0.000 (0.980) | |  | -0.026 (0.968) | 0.098 (1.020) | 0.000 (0.980) | |  | 0.109 (0.995) | -0.029 (0.974) | 0.000 (0.980) | |
| Range | -6.15 - 2.191 | -7.386 - 2.575 | -7.386 - 2.575 | |  | -7.386 - 2.575 | -4.822 - 2.252 | -7.386 - 2.575 | |  | -5.076 - 2.252 | -7.386 - 2.575 | -7.386 - 2.575 | |
| Mean corpuscular volume | | |  | 0.000005 |  |  |  |  | 1 |  |  |  |  | 1 |
| Mean (SD) | -0.036 (0.955) | 0.125 (1.015) | 0.000 (0.971) | |  | -0.005 (0.983) | 0.021 (0.922) | 0.000 (0.971) | |  | 0.001 (0.962) | -0.000 (0.973) | 0.000 (0.971) | |
| Range | -7.343 - 4.665 | -5.246 - 3.931 | -7.343 - 4.665 | |  | -7.343 - 4.665 | -5.246 - 2.647 | -7.343 - 4.665 | |  | -5.246 - 3.292 | -7.343 - 4.665 | -7.343 - 4.665 | |
| Red blood cell count | |  |  | 0.023279 |  |  |  |  | 0.046677 |  |  |  |  | 0.012279 |
| Mean (SD) | 0.035 (0.939) | -0.120 (1.106) | 0.000 (0.981) | |  | -0.024 (0.978) | 0.094 (0.988) | 0.000 (0.981) | |  | 0.110 (0.978) | -0.029 (0.980) | 0.000 (0.981) | |
| Range | -5.862 - 2.818 | -7.334 - 2.734 | -7.334 - 2.818 | |  | -7.334 - 2.818 | -4.405 - 2.631 | -7.334 - 2.818 | |  | -4.405 - 2.631 | -7.334 - 2.818 | -7.334 - 2.818 | |
| Mean platelet volume | |  |  | 0.093474 |  |  |  |  | 1 |  |  |  |  | 1 |
| Mean (SD) | 0.017 (0.867) | -0.059 (1.254) | 0.000 (0.968) | |  | 0.003 (0.941) | -0.013 (1.063) | 0.000 (0.968) | |  | -0.000 (0.999) | 0.000 (0.959) | 0.000 (0.968) | |
| Range | -6.892 - 0.963 | -6.892 - 0.902 | -6.892 - 0.963 | |  | -6.892 - 0.963 | -6.892 - 0.925 | -6.892 - 0.963 | |  | -6.892 - 0.925 | -6.892 - 0.963 | -6.892 - 0.963 | |
| Monocytes % | |  |  | < 1e-06 |  |  |  |  | < 1e-06 |  |  |  |  | < 1e-06 |
| Mean (SD) | 0.057 (0.901) | -0.196 (1.111) | -0.000 (0.957) | |  | 0.070 (0.936) | -0.269 (0.992) | -0.000 (0.957) | |  | -0.316 (0.968) | 0.085 (0.937) | -0.000 (0.957) | |
| Range | -4.1 - 3.152 | -4.1 - 4.962 | -4.1 - 4.962 |  |  | -4.1 - 4.962 | -4.1 - 2.769 | -4.1 - 4.962 |  |  | -4.1 - 2.205 | -4.1 - 4.962 | -4.1 - 4.962 |  |
| Lymphocytes % | |  |  | < 1e-06 |  |  |  |  | < 1e-06 |  |  |  |  | < 1e-06 |
| Mean (SD) | 0.115 (0.916) | -0.398 (1.056) | -0.000 (0.973) | |  | 0.096 (0.935) | -0.369 (1.025) | -0.000 (0.973) | |  | -0.347 (1.022) | 0.093 (0.938) | -0.000 (0.973) | |
| Range | -4.314 - 2.764 | -4.314 - 3.136 | -4.314 - 3.136 | |  | -3.159 - 3.136 | -4.314 - 2.901 | -4.314 - 3.136 | |  | -4.314 - 2.901 | -4.314 - 3.136 | -4.314 - 3.136 | |
| Eosinophils % | |  |  | < 1e-06 |  |  |  |  | < 1e-06 |  |  |  |  | < 1e-06 |
| Mean (SD) | 0.061 (0.986) | -0.211 (0.828) | -0.000 (0.959) | |  | 0.063 (1.002) | -0.240 (0.727) | -0.000 (0.959) | |  | -0.214 (0.749) | 0.057 (1.001) | -0.000 (0.959) | |
| Range | -0.54 - 7.294 | -0.54 - 6.782 | -0.54 - 7.294 | |  | -0.54 - 7.294 | -0.54 - 5.163 | -0.54 - 7.294 | |  | -0.54 - 3.645 | -0.54 - 7.294 | -0.54 - 7.294 | |
| Basophils % |  |  |  | 0.089627 |  |  |  |  | 0.745676 |  |  |  |  | 1 |
| Mean (SD) | 0.025 (0.977) | -0.086 (0.891) | 0.000 (0.959) | |  | 0.021 (0.977) | -0.079 (0.886) | 0.000 (0.959) | |  | -0.036 (0.946) | 0.010 (0.963) | 0.000 (0.959) | |
| Range | -0.399 - 5.122 | -0.399 - 6.011 | -0.399 - 6.011 | |  | -0.399 - 6.011 | -0.399 - 6.011 | -0.399 - 6.011 | |  | -0.399 - 6.011 | -0.399 - 6.011 | -0.399 - 6.011 | |
| Glucose |  |  |  | < 1e-06 |  |  |  |  | < 1e-06 |  |  |  |  | < 1e-06 |
| Mean (SD) | -0.076 (0.937) | 0.265 (1.019) | 0.000 (0.966) | |  | -0.074 (0.951) | 0.282 (0.974) | 0.000 (0.966) | |  | 0.273 (1.003) | -0.073 (0.943) | 0.000 (0.966) | |
| Range | -3.161 - 4.575 | -3.026 - 4.777 | -3.161 - 4.777 | |  | -3.161 - 4.575 | -1.865 - 4.777 | -3.161 - 4.777 | |  | -1.865 - 4.777 | -3.161 - 4.575 | -3.161 - 4.777 | |
| Neutrophils % | |  |  | < 1e-06 |  |  |  |  | < 1e-06 |  |  |  |  | < 1e-06 |
| Mean (SD) | -0.034 (0.800) | 0.118 (1.341) | 0.000 (0.950) | |  | -0.055 (0.963) | 0.212 (0.867) | 0.000 (0.950) | |  | 0.211 (0.861) | -0.057 (0.965) | 0.000 (0.950) | |
| Range | -11.873 - 1.242 | -18.918 - 1.109 | -18.918 - 1.242 | |  | -18.918 - 1.198 | -12.647 - 1.242 | -18.918 - 1.242 | |  | -12.647 - 1.242 | -18.918 - 1.198 | -18.918 - 1.242 | |
| Anion gap |  |  |  | 1 |  |  |  |  | 0.000066 |  |  |  |  | < 1e-06 |
| Mean (SD) | 0.003 (0.938) | -0.011 (1.001) | 0.000 (0.953) | |  | -0.032 (0.948) | 0.122 (0.963) | 0.000 (0.953) | |  | 0.222 (1.011) | -0.060 (0.928) | 0.000 (0.953) | |
| Range | -5.374 - 2.171 | -5.374 - 2.438 | -5.374 - 2.438 | |  | -5.374 - 2.438 | -5.374 - 2.309 | -5.374 - 2.438 | |  | -5.374 - 2.309 | -5.374 - 2.438 | -5.374 - 2.438 | |
| Creatinine |  |  |  | < 1e-06 |  |  |  |  | < 1e-06 |  |  |  |  | 0.000287 |
| Mean (SD) | -0.088 (0.930) | 0.305 (1.025) | -0.000 (0.966) | |  | -0.036 (0.952) | 0.139 (1.004) | -0.000 (0.966) | |  | 0.113 (1.027) | -0.030 (0.946) | -0.000 (0.966) | |
| Range | -2.091 - 6.175 | -1.266 - 5.477 | -2.091 - 6.175 | |  | -1.845 - 6.175 | -2.091 - 5.477 | -2.091 - 6.175 | |  | -2.091 - 5.676 | -1.845 - 6.175 | -2.091 - 6.175 | |
| Sodium |  |  |  | 0.296504 |  |  |  |  | 0.000036 |  |  |  |  | 0.105828 |
| Mean (SD) | -0.001 (1.088) | 0.004 (0.533) | -0.000 (0.991) | |  | 0.032 (0.435) | -0.124 (2.000) | -0.000 (0.991) | |  | -0.101 (2.003) | 0.027 (0.409) | -0.000 (0.991) | |
| Range | -53.983 - 2.599 | -2.837 - 2.247 | -53.983 - 2.599 | |  | -4.631 - 2.331 | -53.983 - 2.599 | -53.983 - 2.599 | |  | -53.983 - 2.599 | -2.48 - 2.279 | -53.983 - 2.599 | |
| Calcium |  |  |  | < 1e-06 |  |  |  |  | < 1e-06 |  |  |  |  | < 1e-06 |
| Mean (SD) | 0.065 (0.966) | -0.226 (0.954) | -0.000 (0.971) | |  | 0.061 (0.990) | -0.234 (0.856) | -0.000 (0.971) | |  | -0.232 (1.381) | 0.062 (0.816) | -0.000 (0.971) | |
| Range | -28.1 - 3.614 | -8.077 - 5.424 | -28.1 - 5.424 | |  | -28.1 - 5.424 | -7.955 - 3.03 | -28.1 - 5.424 | |  | -28.1 - 3.03 | -8.077 - 5.424 | -28.1 - 5.424 | |
| Carbon dioxide | |  |  | 0.007542 |  |  |  |  | 0.001869 |  |  |  |  | < 1e-06 |
| Mean (SD) | 0.034 (0.895) | -0.116 (1.150) | 0.000 (0.960) | |  | 0.035 (0.909) | -0.133 (1.124) | 0.000 (0.960) | |  | -0.322 (1.565) | 0.086 (0.691) | 0.000 (0.960) | |
| Range | -13.35 - 2.588 | -13.35 - 2.08 | -13.35 - 2.588 | |  | -13.35 - 2.08 | -13.35 - 2.588 | -13.35 - 2.588 | |  | -13.35 - 2.08 | -13.35 - 2.588 | -13.35 - 2.588 | |
| Chloride |  |  |  | 1 |  |  |  |  | < 1e-06 |  |  |  |  | < 1e-06 |
| Mean (SD) | -0.010 (1.083) | 0.033 (0.562) | 0.000 (0.991) | |  | 0.030 (0.830) | -0.113 (1.446) | 0.000 (0.991) | |  | -0.110 (1.458) | 0.029 (0.820) | 0.000 (0.991) | |
| Range | -37.708 - 2.333 | -3.685 - 2.151 | -37.708 - 2.333 | |  | -37.708 - 2.313 | -37.708 - 2.333 | -37.708 - 2.333 | |  | -37.708 - 2.333 | -37.708 - 2.313 | -37.708 - 2.333 | |
| Blood urea nitrogen | |  |  | < 1e-06 |  |  |  |  | < 1e-06 |  |  |  |  | 0.000593 |
| Mean (SD) | -0.144 (0.907) | 0.501 (0.981) | -0.000 (0.962) | |  | -0.048 (0.962) | 0.185 (0.940) | -0.000 (0.962) | |  | 0.140 (1.003) | -0.038 (0.948) | -0.000 (0.962) | |
| Range | -4.518 - 3.849 | -4.518 - 3.414 | -4.518 - 3.849 | |  | -4.518 - 3.849 | -2.862 - 3.414 | -4.518 - 3.849 | |  | -2.862 - 3.849 | -4.518 - 3.656 | -4.518 - 3.849 | |
| Potassium |  |  |  | 0.014787 |  |  |  |  | 1 |  |  |  |  | 1 |
| Mean (SD) | -0.016 (0.943) | 0.057 (0.994) | -0.000 (0.955) | |  | -0.006 (0.944) | 0.025 (0.997) | -0.000 (0.955) | |  | 0.033 (0.974) | -0.009 (0.950) | -0.000 (0.955) | |
| Range | -6.432 - 2.594 | -6.432 - 2.791 | -6.432 - 2.791 | |  | -6.432 - 2.594 | -6.432 - 2.791 | -6.432 - 2.791 | |  | -6.432 - 2.791 | -6.432 - 2.594 | -6.432 - 2.791 | |
| Nucleated rbc,absolute | |  |  | < 1e-06 |  |  |  |  | < 1e-06 |  |  |  |  | < 1e-06 |
| Mean (SD) | -0.037 (0.739) | 0.129 (1.565) | 0.000 (0.988) | |  | -0.035 (0.856) | 0.136 (1.374) | 0.000 (0.988) | |  | 0.130 (1.441) | -0.035 (0.823) | 0.000 (0.988) | |
| Range | -0.1 - 25.374 | -0.1 - 29.211 | -0.1 - 29.211 | |  | -0.1 - 29.211 | -0.1 - 19.353 | -0.1 - 29.211 | |  | -0.1 - 29.211 | -0.1 - 25.374 | -0.1 - 29.211 | |
| Rdw-sd |  |  |  | < 1e-06 |  |  |  |  | 1 |  |  |  |  | 1 |
| Mean (SD) | -0.072 (1.016) | 0.250 (0.822) | 0.000 (0.985) | |  | -0.016 (1.029) | 0.060 (0.792) | 0.000 (0.985) | |  | 0.034 (0.801) | -0.009 (1.029) | 0.000 (0.985) | |
| Range | -22.324 - 4.519 | -2.033 - 3.16 | -22.324 - 4.519 | |  | -22.324 - 4.519 | -2.033 - 3.752 | -22.324 - 4.519 | |  | -2.052 - 4.459 | -22.324 - 4.519 | -22.324 - 4.519 | |
| Nucleated red blood cells % | | |  | 0.001303 |  |  |  |  | < 1e-06 |  |  |  |  | < 1e-06 |
| Mean (SD) | -0.040 (0.852) | 0.138 (1.325) | 0.000 (0.980) | |  | -0.046 (0.844) | 0.177 (1.369) | 0.000 (0.980) | |  | 0.178 (1.412) | -0.048 (0.821) | 0.000 (0.980) | |
| Range | -0.148 - 20.492 | -0.148 - 16.923 | -0.148 - 20.492 | |  | -0.148 - 20.492 | -0.148 - 15.739 | -0.148 - 20.492 | |  | -0.148 - 16.923 | -0.148 - 20.492 | -0.148 - 20.492 | |
| Monocytes absolute | |  |  | 1 |  |  |  |  | 0.076315 |  |  |  |  | 0.1956 |
| Mean (SD) | -0.001 (0.829) | 0.003 (1.067) | -0.000 (0.888) | |  | 0.020 (0.882) | -0.078 (0.907) | -0.000 (0.888) | |  | -0.068 (0.921) | 0.018 (0.878) | -0.000 (0.888) | |
| Range | -2.119 - 4.125 | -2.119 - 14.376 | -2.119 - 14.376 | |  | -2.119 - 14.376 | -2.119 - 3.86 | -2.119 - 14.376 | |  | -2.119 - 3.86 | -2.119 - 14.376 | -2.119 - 14.376 | |
| Lymphocytes absolute | |  |  | < 1e-06 |  |  |  |  | < 1e-06 |  |  |  |  | 0.000035 |
| Mean (SD) | 0.058 (0.853) | -0.200 (0.960) | -0.000 (0.884) | |  | 0.042 (0.862) | -0.161 (0.948) | -0.000 (0.884) | |  | -0.104 (0.955) | 0.028 (0.862) | -0.000 (0.884) | |
| Range | -1.879 - 3.652 | -1.879 - 7.457 | -1.879 - 7.457 | |  | -1.879 - 6.031 | -1.879 - 7.457 | -1.879 - 7.457 | |  | -1.879 - 7.457 | -1.879 - 6.031 | -1.879 - 7.457 | |
| Eosinophils, absolute | |  |  | < 1e-06 |  |  |  |  | < 1e-06 |  |  |  |  | 0.000807 |
| Mean (SD) | 0.037 (0.940) | -0.128 (0.899) | 0.000 (0.934) | |  | 0.037 (0.967) | -0.141 (0.778) | 0.000 (0.934) | |  | -0.119 (0.750) | 0.032 (0.975) | 0.000 (0.934) | |
| Range | -0.424 - 14.025 | -0.424 - 10.696 | -0.424 - 14.025 | |  | -0.424 - 14.025 | -0.424 - 10.696 | -0.424 - 14.025 | |  | -0.424 - 7.231 | -0.424 - 14.025 | -0.424 - 14.025 | |
| Basophils absolute | |  |  | 1 |  |  |  |  | 1 |  |  |  |  | 0.000645 |
| Mean (SD) | -0.008 (0.882) | 0.026 (0.913) | 0.000 (0.889) | |  | -0.014 (0.872) | 0.054 (0.950) | 0.000 (0.889) | |  | 0.108 (1.011) | -0.029 (0.851) | 0.000 (0.889) | |
| Range | -0.353 - 5.633 | -0.353 - 5.633 | -0.353 - 5.633 | |  | -0.353 - 5.633 | -0.353 - 5.633 | -0.353 - 5.633 | |  | -0.353 - 5.633 | -0.353 - 3.36 | -0.353 - 5.633 | |
| Neutrophils absolute | |  |  | < 1e-06 |  |  |  |  | < 1e-06 |  |  |  |  | < 1e-06 |
| Mean (SD) | -0.048 (0.897) | 0.166 (1.079) | -0.000 (0.945) | |  | -0.056 (0.929) | 0.213 (0.974) | -0.000 (0.945) | |  | 0.236 (0.997) | -0.063 (0.920) | -0.000 (0.945) | |
| Range | -3.303 - 3.085 | -3.869 - 3.375 | -3.869 - 3.375 | |  | -3.869 - 3.085 | -3.542 - 3.375 | -3.869 - 3.375 | |  | -3.542 - 3.375 | -3.869 - 3.085 | -3.869 - 3.375 | |
| Albumin |  |  |  | < 1e-06 |  |  |  |  | 0.00842 |  |  |  |  | 1 |
| Mean (SD) | 0.095 (0.876) | -0.328 (1.024) | 0.000 (0.928) | |  | 0.030 (0.924) | -0.114 (0.936) | 0.000 (0.928) | |  | -0.030 (0.927) | 0.008 (0.928) | 0.000 (0.928) | |
| Range | -4.537 - 2.563 | -5.159 - 2.457 | -5.159 - 2.563 | |  | -5.159 - 2.563 | -4.246 - 1.901 | -5.159 - 2.563 | |  | -4.537 - 1.901 | -5.159 - 2.563 | -5.159 - 2.563 | |
| Alt |  |  |  | 1 |  |  |  |  | < 1e-06 |  |  |  |  | 0.000003 |
| Mean (SD) | -0.017 (0.913) | 0.060 (0.875) | -0.000 (0.905) | |  | -0.048 (0.905) | 0.182 (0.879) | -0.000 (0.905) | |  | 0.167 (0.944) | -0.045 (0.889) | -0.000 (0.905) | |
| Range | -4.747 - 6.554 | -4.747 - 4.833 | -4.747 - 6.554 | |  | -4.747 - 6.554 | -2.16 - 4.833 | -4.747 - 6.554 | |  | -4.747 - 6.554 | -4.747 - 4.44 | -4.747 - 6.554 | |
| Bilirubin total | |  |  | 0.009987 |  |  |  |  | 0.27583 |  |  |  |  | 0.023831 |
| Mean (SD) | -0.038 (0.864) | 0.133 (1.087) | -0.000 (0.921) | |  | -0.023 (0.899) | 0.089 (1.000) | -0.000 (0.921) | |  | 0.146 (1.186) | -0.039 (0.833) | -0.000 (0.921) | |
| Range | -1.906 - 11.182 | -1.906 - 8.975 | -1.906 - 11.182 | |  | -1.906 - 11.182 | -1.906 - 8.975 | -1.906 - 11.182 | |  | -1.906 - 11.182 | -1.537 - 8.477 | -1.906 - 11.182 | |
| Ast |  |  |  | < 1e-06 |  |  |  |  | < 1e-06 |  |  |  |  | < 1e-06 |
| Mean (SD) | -0.052 (0.871) | 0.181 (1.047) | 0.000 (0.919) | |  | -0.057 (0.888) | 0.217 (0.999) | 0.000 (0.919) | |  | 0.193 (1.012) | -0.052 (0.885) | 0.000 (0.919) | |
| Range | -4.388 - 5.58 | -4.388 - 4.344 | -4.388 - 5.58 | |  | -4.388 - 5.58 | -4.388 - 4.344 | -4.388 - 5.58 | |  | -4.388 - 5.58 | -4.388 - 4.344 | -4.388 - 5.58 | |
| Bilirubin direct | |  |  | 0.015298 |  |  |  |  | 0.000297 |  |  |  |  | < 1e-06 |
| Mean (SD) | -0.042 (0.830) | 0.146 (1.182) | 0.000 (0.924) | |  | -0.035 (0.878) | 0.133 (1.071) | 0.000 (0.924) | |  | 0.227 (1.319) | -0.061 (0.774) | 0.000 (0.924) | |
| Range | -1.253 - 15.931 | -1.253 - 11.065 | -1.253 - 15.931 | |  | -1.253 - 15.931 | -1.253 - 11.065 | -1.253 - 15.931 | |  | -1.253 - 15.931 | -1.253 - 11.065 | -1.253 - 15.931 | |
| Protein, total | |  |  | 0.000679 |  |  |  |  | 0.037153 |  |  |  |  | 0.023326 |
| Mean (SD) | 0.042 (0.878) | -0.145 (1.021) | 0.000 (0.915) | |  | 0.026 (0.910) | -0.100 (0.927) | 0.000 (0.915) | |  | -0.109 (0.997) | 0.029 (0.889) | 0.000 (0.915) | |
| Range | -6.924 - 3.883 | -7.048 - 3.212 | -7.048 - 3.883 | |  | -6.924 - 3.883 | -7.048 - 2.631 | -7.048 - 3.883 | |  | -7.048 - 3.212 | -4.632 - 3.883 | -7.048 - 3.883 | |
| Alkaline phosphatase | |  |  | 1 |  |  |  |  | 1 |  |  |  |  | 1 |
| Mean (SD) | -0.006 (0.908) | 0.022 (0.899) | -0.000 (0.906) | |  | 0.009 (0.912) | -0.034 (0.882) | -0.000 (0.906) | |  | 0.017 (0.916) | -0.005 (0.903) | -0.000 (0.906) | |
| Range | -9.111 - 5.87 | -2.268 - 4.327 | -9.111 - 5.87 | |  | -9.111 - 5.87 | -2.268 - 4.096 | -9.111 - 5.87 | |  | -2.268 - 4.711 | -9.111 - 5.87 | -9.111 - 5.87 | |
| Troponin i |  |  |  | < 1e-06 |  |  |  |  | 0.010543 |  |  |  |  | 0.000101 |
| Mean (SD) | -0.059 (0.771) | 0.206 (1.186) | 0.000 (0.887) | |  | -0.030 (0.818) | 0.114 (1.108) | 0.000 (0.887) | |  | 0.178 (1.422) | -0.048 (0.668) | 0.000 (0.887) | |
| Range | -0.268 - 15.421 | -0.268 - 10.578 | -0.268 - 15.421 | |  | -0.268 - 15.421 | -0.268 - 13.543 | -0.268 - 15.421 | |  | -0.268 - 15.421 | -0.268 - 11.09 | -0.268 - 15.421 | |
| Granulocytes, immature % | | |  | < 1e-06 |  |  |  |  | < 1e-06 |  |  |  |  | < 1e-06 |
| Mean (SD) | -0.070 (0.897) | 0.245 (0.995) | 0.000 (0.929) | |  | -0.067 (0.889) | 0.257 (1.027) | 0.000 (0.929) | |  | 0.252 (1.022) | -0.067 (0.890) | 0.000 (0.929) | |
| Range | -1.028 - 3.194 | -1.028 - 4.574 | -1.028 - 4.574 | |  | -1.028 - 3.194 | -1.028 - 4.574 | -1.028 - 4.574 | |  | -1.028 - 4.574 | -1.028 - 3.194 | -1.028 - 4.574 | |
| Granulocytes immature , absolute | | |  | < 1e-06 |  |  |  |  | < 1e-06 |  |  |  |  | < 1e-06 |
| Mean (SD) | -0.074 (0.790) | 0.256 (1.160) | -0.000 (0.897) | |  | -0.072 (0.799) | 0.277 (1.159) | -0.000 (0.897) | |  | 0.297 (1.158) | -0.080 (0.794) | -0.000 (0.897) | |
| Range | -0.625 - 5.464 | -0.625 - 6.301 | -0.625 - 6.301 | |  | -0.625 - 5.795 | -0.625 - 6.301 | -0.625 - 6.301 | |  | -0.625 - 6.301 | -0.625 - 5.802 | -0.625 - 6.301 | |
| Ldh |  |  |  | < 1e-06 |  |  |  |  | < 1e-06 |  |  |  |  | < 1e-06 |
| Mean (SD) | -0.039 (0.843) | 0.135 (0.848) | -0.000 (0.847) | |  | -0.035 (0.831) | 0.134 (0.895) | -0.000 (0.847) | |  | 0.077 (0.946) | -0.021 (0.818) | -0.000 (0.847) | |
| Range | -3.32 - 1.368 | -3.32 - 1.499 | -3.32 - 1.499 | |  | -3.32 - 1.368 | -3.32 - 1.499 | -3.32 - 1.499 | |  | -3.32 - 1.499 | -3.32 - 1.483 | -3.32 - 1.499 | |
| C-reactive protein | |  |  | < 1e-06 |  |  |  |  | < 1e-06 |  |  |  |  | < 1e-06 |
| Mean (SD) | -0.086 (0.887) | 0.297 (0.705) | 0.000 (0.865) | |  | -0.090 (0.886) | 0.346 (0.675) | 0.000 (0.865) | |  | 0.292 (0.706) | -0.078 (0.886) | 0.000 (0.865) | |
| Range | -3.407 - 1.429 | -3.407 - 1.672 | -3.407 - 1.672 | |  | -3.407 - 1.672 | -3.407 - 1.501 | -3.407 - 1.672 | |  | -3.407 - 1.501 | -3.407 - 1.672 | -3.407 - 1.672 | |
| Prothrombin time | |  |  | < 1e-06 |  |  |  |  | 0.000033 |  |  |  |  | 0.005524 |
| Mean (SD) | -0.053 (0.787) | 0.185 (0.979) | 0.000 (0.839) | |  | -0.032 (0.801) | 0.123 (0.962) | 0.000 (0.839) | |  | 0.080 (0.822) | -0.022 (0.843) | 0.000 (0.839) | |
| Range | -10.887 - 9.976 | -2.362 - 9.221 | -10.887 - 9.976 | |  | -10.887 - 9.976 | -2.679 - 9.221 | -10.887 - 9.976 | |  | -2.679 - 7.088 | -10.887 - 9.976 | -10.887 - 9.976 | |
| Inr |  |  |  | < 1e-06 |  |  |  |  | 0.00007 |  |  |  |  | 0.020877 |
| Mean (SD) | -0.051 (0.780) | 0.177 (1.041) | 0.000 (0.851) | |  | -0.030 (0.797) | 0.113 (1.024) | 0.000 (0.851) | |  | 0.062 (0.815) | -0.017 (0.860) | 0.000 (0.851) | |
| Range | -4.978 - 12.245 | -1.009 - 11.181 | -4.978 - 12.245 | |  | -4.978 - 12.245 | -2.033 - 11.181 | -4.978 - 12.245 | |  | -2.033 - 8.249 | -4.978 - 12.245 | -4.978 - 12.245 | |
| D-dimer, quantitative | |  |  | < 1e-06 |  |  |  |  | 0.070659 |  |  |  |  | 0.000011 |
| Mean (SD) | -0.039 (0.819) | 0.136 (0.838) | 0.000 (0.827) | |  | -0.019 (0.829) | 0.073 (0.813) | 0.000 (0.827) | |  | 0.130 (0.776) | -0.035 (0.836) | 0.000 (0.827) | |
| Range | -2.854 - 2.504 | -2.854 - 2.513 | -2.854 - 2.513 | |  | -2.854 - 2.513 | -2.854 - 2.366 | -2.854 - 2.513 | |  | -2.854 - 2.513 | -2.854 - 2.504 | -2.854 - 2.513 | |
| Ferritin |  |  |  | < 1e-06 |  |  |  |  | < 1e-06 |  |  |  |  | < 1e-06 |
| Mean (SD) | -0.056 (0.864) | 0.195 (0.778) | 0.000 (0.852) | |  | -0.058 (0.858) | 0.224 (0.791) | 0.000 (0.852) | |  | 0.211 (0.810) | -0.057 (0.854) | 0.000 (0.852) | |
| Range | -4.382 - 2.793 | -4.382 - 2.556 | -4.382 - 2.793 | |  | -4.382 - 2.793 | -4.382 - 2.556 | -4.382 - 2.793 | |  | -4.382 - 2.556 | -4.382 - 2.793 | -4.382 - 2.793 | |
| Procalcitonin | |  |  | < 1e-06 |  |  |  |  | < 1e-06 |  |  |  |  | < 1e-06 |
| Mean (SD) | -0.069 (0.683) | 0.238 (1.072) | -0.000 (0.797) | |  | -0.043 (0.735) | 0.163 (0.984) | -0.000 (0.797) | |  | 0.175 (1.002) | -0.047 (0.726) | -0.000 (0.797) | |
| Range | -0.569 - 9.354 | -0.569 - 8.536 | -0.569 - 9.354 | |  | -0.569 - 9.246 | -0.569 - 9.354 | -0.569 - 9.354 | |  | -0.569 - 8.536 | -0.569 - 9.354 | -0.569 - 9.354 | |
