## Supplemental Table S2 for "Predictive modeling of morbidity and mortality in COVID-19 hospitalized patients and its clinical implications"

2. XGboost parameter settings for each trained model.

| XGBoost | without Labs |  |  |  |  |  |  |  |
| --- | --- | --- | --- | --- | --- | --- | --- | --- |
|  |  | Day 0 |  |  |  | Day -1 |  |  |
|  |  | Deceased | Ventilated | ICU |  | Deceased | Ventilated | ICU |
|  | eta | 0.01 | 0.01 | 0.01 |  | 0.01 | 0.01 | 0.01 |
|  | Max depth | 3 | 3 | 2 |  | 3 | 4 | 4 |
|  | Gamma | 0.4 | 0.6 | 0.2 |  | 0.2 | 1 | 0.8 |
|  | Min child weight | 0.6 | 0.2 | 0.8 |  | 0.6 | 0.2 | 0.2 |
|  | with Labs | (Averaged across 5 individually built models) | | | |  |  |  |
|  |  | Day 0 |  |  |  |  |  |  |
|  |  | Deceased | Ventilated | ICU |  |  |  |  |
|  | eta | 0.01 | 0.01 | 0.028 |  |  |  |  |
|  | Max depth | 3.8 | 2.8 | 3.2 |  |  |  |  |
|  | Gamma | 0.4 | 0.64 | 0.36 |  |  |  |  |
|  | Min child weight | 0.44 | 0.56 | 0.48 |  |  |  |  |
|  | Subset by Age | (Averaged across 5 individually built models) | | | |  |  |  |
|  |  | Day 0 |  |  |  |  |  |  |
|  |  | Deceased |  | Ventilated |  | ICU |  |  |
|  |  | >=65 | 18-64 | >=65 | 18-64 | >=65 | 18-64 |  |
|  | eta | 0.01 | 0.1 | 0.01 | 0.01 | 0.01 | 0.028 |  |
|  | Max depth | 3.6 | 6.6 | 2.2 | 2.8 | 2.2 | 3.4 |  |
|  | Gamma | 0.68 | 0.48 | 0.48 | 0.6 | 0.8 | 0.52 |  |
|  | Min child weight | 0.32 | 0.64 | 0.6 | 0.44 | 0.52 | 0.84 |  |
