## Supplemental Table S3 for "Predictive modeling of morbidity and mortality in COVID-19 hospitalized patients and its clinical implications"

3. Description of features utilized for time-series modeling.

| **vital sign features** | |  |  | **lab results features** |  |  |  | **all features** |  |  |
| --- | --- | --- | --- | --- | --- | --- | --- | --- | --- | --- |
| **variable** | **type** | **standardized** |  | **variable** | **type** | **standardized** |  | **variable** | **type** | **standardized** |
| progression | numeric | no |  | progression | numeric | no |  | sex | binary | no |
| pulse | numeric | yes |  | pulse | numeric | yes |  | diabetes | binary | no |
| spo2 | numeric | yes |  | spo2 | numeric | yes |  | comorbids | binary | no |
| temp | numeric | yes |  | temp | numeric | yes |  | age_0-17 | binary | no |
| resp | numeric | yes |  | resp | numeric | yes |  | age_18-44 | binary | no |
| sbp | numeric | yes |  | sbp | numeric | yes |  | age_45-64 | binary | no |
| dbp | numeric | yes |  | dbp | numeric | yes |  | age_65-74 | binary | no |
|  |  |  |  | WHITE.BLOOD.CELL.COUNT | numeric | yes |  | age_75+ | binary | no |
|  |  |  |  | MEAN.CORPUSCULAR.HEMOGLOBIN | numeric | yes |  | age_NULL | binary | no |
|  |  |  |  | MEAN.CORPUSCULAR.HEMOGLOBIN.CONC | numeric | yes |  | progression | numeric | no |
|  |  |  |  | PLATELET.COUNT | numeric | yes |  | pulse | numeric | yes |
|  |  |  |  | RDW.CV | numeric | yes |  | spo2 | numeric | yes |
|  |  |  |  | HEMATOCRIT | numeric | yes |  | temp | numeric | yes |
|  |  |  |  | HEMOGLOBIN | numeric | yes |  | resp | numeric | yes |
|  |  |  |  | MEAN.CORPUSCULAR.VOLUME | numeric | yes |  | sbp | numeric | yes |
|  |  |  |  | RED.BLOOD.CELL.COUNT | numeric | yes |  | dbp | numeric | yes |
|  |  |  |  | MEAN.PLATELET.VOLUME | numeric | yes |  | WHITE.BLOOD.CELL.COUNT | numeric | yes |
|  |  |  |  | MONOCYTES.. | numeric | yes |  | MEAN.CORPUSCULAR.HEMOGLOBIN | numeric | yes |
|  |  |  |  | LYMPHOCYTES.. | numeric | yes |  | MEAN.CORPUSCULAR.HEMOGLOBIN.CONC | numeric | yes |
|  |  |  |  | EOSINOPHILS.. | numeric | yes |  | PLATELET.COUNT | numeric | yes |
|  |  |  |  | BASOPHILS.. | numeric | yes |  | RDW.CV | numeric | yes |
|  |  |  |  | GLUCOSE | numeric | yes |  | HEMATOCRIT | numeric | yes |
|  |  |  |  | NEUTROPHILS.. | numeric | yes |  | HEMOGLOBIN | numeric | yes |
|  |  |  |  | ANION.GAP | numeric | yes |  | MEAN.CORPUSCULAR.VOLUME | numeric | yes |
|  |  |  |  | CREATININE | numeric | yes |  | RED.BLOOD.CELL.COUNT | numeric | yes |
|  |  |  |  | SODIUM | numeric | yes |  | MEAN.PLATELET.VOLUME | numeric | yes |
|  |  |  |  | CALCIUM | numeric | yes |  | MONOCYTES.. | numeric | yes |
|  |  |  |  | CARBON.DIOXIDE | numeric | yes |  | LYMPHOCYTES.. | numeric | yes |
|  |  |  |  | CHLORIDE | numeric | yes |  | EOSINOPHILS.. | numeric | yes |
|  |  |  |  | BLOOD.UREA.NITROGEN | numeric | yes |  | BASOPHILS.. | numeric | yes |
|  |  |  |  | POTASSIUM | numeric | yes |  | GLUCOSE | numeric | yes |
|  |  |  |  | ALBUMIN | numeric | yes |  | NEUTROPHILS.. | numeric | yes |
|  |  |  |  | ALT | numeric | yes |  | ANION.GAP | numeric | yes |
|  |  |  |  | BILIRUBIN.TOTAL | numeric | yes |  | CREATININE | numeric | yes |
|  |  |  |  | AST | numeric | yes |  | SODIUM | numeric | yes |
|  |  |  |  | BILIRUBIN.DIRECT | numeric | yes |  | CALCIUM | numeric | yes |
|  |  |  |  | PROTEIN..TOTAL | numeric | yes |  | CARBON.DIOXIDE | numeric | yes |
|  |  |  |  | ALKALINE.PHOSPHATASE | numeric | yes |  | CHLORIDE | numeric | yes |
|  |  |  |  | TROPONIN.I | numeric | yes |  | BLOOD.UREA.NITROGEN | numeric | yes |
|  |  |  |  | GRANULOCYTES..IMMATURE.. | numeric | yes |  | POTASSIUM | numeric | yes |
|  |  |  |  | LDH | numeric | yes |  | ALBUMIN | numeric | yes |
|  |  |  |  | C.REACTIVE.PROTEIN | numeric | yes |  | ALT | numeric | yes |
|  |  |  |  | PROTHROMBIN.TIME | numeric | yes |  | BILIRUBIN.TOTAL | numeric | yes |
|  |  |  |  | INR | numeric | yes |  | AST | numeric | yes |
|  |  |  |  | D.DIMER..QUANTITATIVE | numeric | yes |  | BILIRUBIN.DIRECT | numeric | yes |
|  |  |  |  | FERRITIN | numeric | yes |  | PROTEIN..TOTAL | numeric | yes |
|  |  |  |  | PROCALCITONIN | numeric | yes |  | ALKALINE.PHOSPHATASE | numeric | yes |
|  |  |  |  |  |  |  |  | TROPONIN.I | numeric | yes |
|  |  |  |  |  |  |  |  | GRANULOCYTES..IMMATURE.. | numeric | yes |
|  |  |  |  |  |  |  |  | LDH | numeric | yes |
|  |  |  |  |  |  |  |  | C.REACTIVE.PROTEIN | numeric | yes |
|  |  |  |  |  |  |  |  | PROTHROMBIN.TIME | numeric | yes |
|  |  |  |  |  |  |  |  | INR | numeric | yes |
|  |  |  |  |  |  |  |  | D.DIMER..QUANTITATIVE | numeric | yes |
|  |  |  |  |  |  |  |  | FERRITIN | numeric | yes |
|  |  |  |  |  |  |  |  | PROCALCITONIN | numeric | yes |
