## Supplemental Figure 1 for "Predictive modeling of morbidity and mortality in COVID-19 hospitalized patients and its clinical implications"

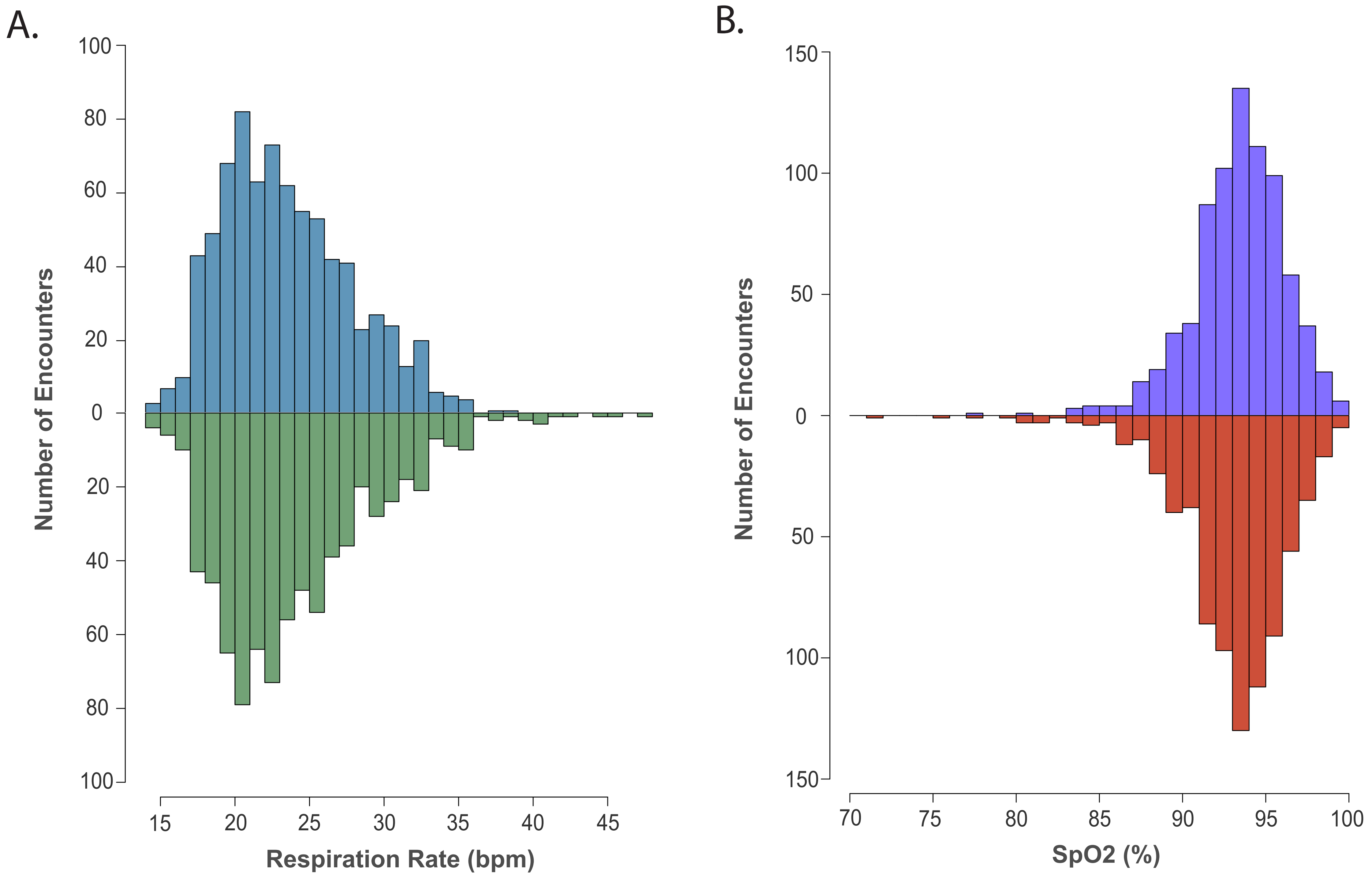


| Respiration Rate (bpm) | Mean (bpm) | SD (bpm) |
|---|---|---|
| All | 23.63 | 4.35 |
| Filtered Subset | 24.12 | 5.08 |

| SpO2 (%) | Mean (%) | SD (%) |
|---|---|---|
| All | 93.47 | 2.78 |
| Filtered Subset | 93.16 | 3.23 |
